# Improved water access may not reduce women’s time burdens: Evidence from Kenya and Honduras

**DOI:** 10.64898/2026.06.03.26354805

**Authors:** Sheela Sinharoy, Thea Mink, Emily Ogutu, Madeleine Patrick, Maria del Carmen Alvarez Nuncio, Margot V. Bolaños-Gamez, Harriet Oglesby, Christopher P. Ngo, Sandra Antonio, Edwin Medina Lopez, Peter Mwangi, Paul Ruto, Peter Koome, Petronilla Andiba Otuya, Rohin Otieno Onyango, Bethany Caruso

## Abstract

Women’s disproportionate responsibility for unpaid domestic and care work, including water collection, remains a barrier to gender equality globally and may constrain women’s ability to engage in income-generating activities. We compared women’s and men’s time use in rural Kenya and Honduras and assessed whether women’s time spent on water collection and income-generating activities differed between communities that had or had not received an improved water source from World Vision. We also examined the measurement of time-use agency among women and men.

In-person surveys were conducted in July–August 2024 with 95 participants (48 women, 47 men) in six Kenyan communities and 102 participants (53 women, 49 men) in six Honduran communities. Surveys included a 24-hour time-use recall module and items on time-use agency. Analyses compared time use by gender and by community intervention status (improved vs. not yet improved water supply), and confirmatory factor analysis assessed the validity of the time-use agency measure.

Women in both study sites spent substantially more time than men on unpaid domestic and care work activities, including cooking, cleaning, laundry, and caregiving. In Kenya, women also spent significantly more time collecting water. Men spent more time sleeping (Kenya), on paid work (Honduras), unpaid agricultural work (both settings), and traveling (both settings). Across both countries, there were no significant differences between intervention and comparison communities in women’s time spent on water collection or income-generating activities. In Kenya, most respondents reported high influence over their time, and six items showed strong validity for measuring instrumental time-use agency.

Women’s time burdens remained high even in communities that had received improved water sources, including at the household level. Our results suggest that more transformative water infrastructure, combined with interventions that address gendered social norms, may be needed to meaningfully reduce women’s domestic work burden and support their economic empowerment.

## INTRODUCTION

Measuring women’s unpaid care and domestic work is widely recognized as a pressing global priority, yet data collection efforts remain insufficient. Sustainable Development Goal (SDG) 5 seeks to “Achieve gender equality and empower all women and girls” and specifies Target 5.4 which aims to “Recognize and value unpaid care and domestic work through the provision of public services, infrastructure and social protection policies and the promotion of shared responsibility within the household and the family as nationally appropriate”.[1] However, of the 169 SDG targets, Target 5.4 is one of only 34 that are not tracked sufficiently for regularly reporting due to a lack of available global data for its associated indicator (5.4.1: “Proportion of time spent on unpaid domestic and care work, by sex, age and location”).[2, 3] Target 5.4 is considered a ‘Tier II’ target, meaning that it has a conceptually clear indicator with internationally established standards for data collection, but countries do not regularly produce data (compared to Tier I, in which at least 50% of countries report data).[2] Therefore, large gaps exist in our knowledge of how women and men spend their time across global settings.

From the evidence that does exist, we know that women spend disproportionate time on unpaid care and domestic work and that, especially in rural areas of low- and middle-income countries, this work often includes water collection.[4] The large burden of time spent on unpaid care and domestic work has important implications for women’s ability to engage in other activities, including income-generating activities, other productive work, health-promoting behaviors (including rest and leisure), or health-seeking behaviors.[5, 6] For example, our previous research observed that women in selected rural areas of Guatemala, Honduras, Kenya, and Zimbabwe spent, on average, 82 minutes per trip to collect water (range of means across countries: 38 minutes (Honduras)-164 minutes (Kenya)).[7] Women described how water collection negatively affected their health and wellbeing [7], demanded both cognitive and emotional labor, and limited their available time to engage in income-generating activities.[8, 9] Of note, those results focused only on water collection and did not take into consideration the many other activities that women perform each day, which would further add to their time burden of unpaid and domestic work.

In recognition of these time burdens, it has been argued that provision of services such as water, sanitation, electricity, and transport has the potential to decrease unpaid work time, particularly for women.[6, 10, 11] This hypothesis has been examined in several studies, with varying results. A systematic review and meta-analysis of the effect of water supply interventions on time spent accessing water found a statistically significant reduction in time spent traveling and accessing water (d = −0.21; 95% CI = −0.29, −0.13; 25 estimates) following intervention.[12] Results of the meta-analysis indicated that the travel time saved (in minutes per trip) following water supply interventions was eight minutes on average (95% CI = −13, −2; 11 estimates).[12] In the few studies that measured how this time was reallocated, there was no clear pattern and no significant increase or decrease in time spent on other activities such as paid work or leisure.[12] Therefore, while the current evidence indicates that water supply interventions are effective for reducing time spent on water collection to some degree, it is not clear that they are effective in decreasing unpaid work time overall or increasing time spent on other types of activities.

One challenge in measuring and comparing these outcomes is the diversity of existing methods of data collection on time use. Among national-level surveys implemented by governments’ national statistical offices and similar entities, a lack of harmonization of approaches remains a key challenge.[4] In 2024, the UN Statistical Commission released a *Guide to producing statistics on time use*, to support harmonization and comparability.[13] Among other national-level data collection efforts, the Demographic and Health Survey (DHS) Program and other global household surveys often include questions on time use, but they typically focus only on a few specific activities. For example, the DHS includes questions to assess time spent on water collection, which are used by the WHO/UNICEF Joint Monitoring Programme for Water Supply, Sanitation and Hygiene (JMP) to monitor and report on SDG Target 6.1. However, assessing singular activities, as in the DHS, does not capture an individual’s complete workload or inequalities in workloads within households. Existing tools that seek to assess an individual’s complete workload do exist. For example, the Women’s Empowerment in Agriculture Index (WEAI) family of survey instruments, which have also been used at the national level, includes a time allocation module to assess time use (including paid and unpaid work) over a 24-hour period. However, the WEAI does not include a separate activity code for water collection, only a broad code for domestic work under which water collection would be included among other domestic work activities. Therefore, data from these different survey instruments are not comparable and often lack details on specific types of unpaid and domestic work.

A different type of challenge is that while provision of services may save time for women, women may have little control over this time, due to gendered social norms that dictate expected roles and responsibilities.[8, 14] Because of these norms, time savings from water collection may simply be reallocated to other unpaid and domestic care work or may be reallocated in ways that generate time for other family members.[15] For example, in a study in Ghana, a rural water supply project led to time savings for women and men, which women, more than men, reallocated to create time for others (for example, by taking on additional chores to allow men to spend more time in the fields).[16] In many contexts, women’s time may be considered a communal or collective resource that is made available to benefit others rather than being fully under their own control.[17, 18] For this reason, not only time use but the more recently developed concept of time-use agency – defined as “an individual’s confidence and ability to make and act upon strategic choices about how to allocate one’s time”[14] -- are both considered to be important for women’s empowerment.

### Study Objective

As described above, large gaps exist in our knowledge of (a) how women and men spend their time across global settings; (b) whether provision of improved water supply can meaningfully reduce women’s time burden on unpaid and domestic work and, if so, how any time savings may be reallocated; and (c) whether women have control over their time, which would be a prerequisite for reallocating time that is saved through water supply interventions. To address these evidence gaps, our study had three corresponding aims, for which we collected data within the context of World Vision programming in rural areas of Kenya and Honduras. First, we aimed to measure, report, and compare time use among women and men in each of our study settings, with a particular focus on time spent on water collection and income-generating activities. Second, among women in each setting, we aimed to compare time spent on water collection and income-generating activities between communities that had or had not received water infrastructure through World Vision programming. Third, we aimed to compare aspects of time-use agency between women and men and assess the construct validity of a scale to measure time-use agency in new settings.

## METHODS

### Study design

The present study involved cross-sectional surveys among women and men in rural areas of Kenya and Honduras where World Vision is delivering its *Strong Women, Strong World: Beyond Access* (SWSW) programming. The SWSW program aims to increase women’s empowerment and well-being through a multi-component intervention that integrates water improvements and economic empowerment training and support. The SWSW program theorizes that the provision of improved water services will lead to increased access to water, thereby reducing women’s time and energy expenditures and enabling them to use water, time, and energy for other purposes, including income-generating activities. To better understand women’s and men’s time allocation, including for water collection and economic (income-generating) activities, Emory University worked with World Vision USA and local World Vision offices in Honduras and Kenya to design and conduct the present study. Additional local learning partners who contributed research expertise and led data collection activities were the Edwin Roldan Medina Lopez Research Group in Honduras and St. Paul’s University in Kenya.

### Study setting

Kenya data collection occurred in rural areas of Samburu and Isiolo counties. The region is typically arid and semi-arid. However, during data collection for the present study (July-August 2024), the study communities were experiencing unseasonably heavy rainfall. Of the total population in Kenya, 66% have at least basic water service (56% rural, 89% urban); in Isiolo county, 72% have at least basic water service and in Samburu county only 29% have at least basic water service.[19, 20] Honduras data collection occurred in rural and peri-urban areas in Danli and Teupasenti municipalities. The study areas are forested and have a tropical climate; data collection occurred during the rainy season (July-August 2024). Of the total population, 96% have at least basic water service (90% rural, > 99% urban).[20]

### Participant selection and recruitment

The present study was nested within a larger study of women’s water collection experiences. As such, sample size and participant selection procedures were also nested within the larger study and are described in detail elsewhere.[7, 21] Briefly, six communities were purposively selected per country in collaboration with World Vision. In Kenya, three of the six communities (hereafter referred to as ‘intervention communities’) had received an improved water source (specifically, a water kiosk connected to a water tank) through World Vision programming, while the other three communities (hereafter, ‘comparison communities’) had not yet received the water infrastructure from World Vision but were slated to do so later in 2024 or 2025. Two of the three intervention communities had participated in research activities the prior year.[7] In Honduras, two of the six communities had received an improved water source (specifically, household piped water), while the other four had not. Both intervention communities and one of the comparison communities had participated in research activities the prior year, while the remaining three comparison communities had not been included in previous research.

To select individual participants, as part of the larger study on water collection experiences, field teams worked with community leaders to recruit eight women per community for data collection. World Vision staff did not participate in recruitment of study participants. Women were eligible if they were aged 18 years or older, responsible for collecting water for household needs, and lived in eligible communities. Exclusion criteria were physical disabilities or barriers that would prevent them from participating in the larger research study and responding to survey questions. In communities where we had previously collected data, individuals who had participated in the previous round of research and who had shared their contact information were purposively recruited and invited to participate again. For comparison communities that were not part of the previous research, the teams used a convenience sampling approach to recruit participants. While the primary focus was on women, we also included men in the study for a broader understanding of time use in the selected communities. Household dyads were preferred, meaning that when a married woman was recruited and enrolled, an effort was made to recruit her husband. When single or widowed women were recruited, a single or widowed man of a similar age from the same community was sought to be recruited, with the aim of recruiting and enrolling an equal number of women and men.

### Data collection tools and procedures

We adapted the project-level WEAI (pro-WEAI) time allocation module to collect data on time use.[22, 23] The pro-WEAI module uses an open-recall approach, in which an interviewer asks the participant to report all activities undertaken during the previous day in chronological order. The interviewer records activities using an activity list, assigning them to 15-minute time slots. We adapted the activity categories in three ways. First, the pro-WEAI module has one category for “domestic work” that includes fetching water and fuel; for the purposes of our study, we created two mutually exclusive categories for water collection and other domestic work. Second, the pro-WEAI, being focused on agriculture, includes six different activity categories for agricultural work. We reduced this to one category for agricultural work, including farming, livestock rearing, and, in Honduras only, fishing. Third, because World Vision programming in Kenya and Honduras is focused on women’s economic empowerment, we separated several activities into two codes to indicate if the activity was mainly for the self or for household use and consumption, or if it was mainly to enable sale or trade. For example, we had one category for agricultural activities, but we split this into two activity codes to represent (a) agricultural activities that were mainly for the household’s own consumption versus (b) agricultural activities that were mainly for sale or trade.

We also adapted and used a time-use agency scale that had originally been developed and tested among a population of rural women and men within a project in Ghana.[24] The original time-use agency scale consisted of 30 items: 8 items on intrinsic agency (including the concepts of self-efficacy and critical consciousness) and 22 items on instrumental agency (including decision-making and voice). To reduce time and participant cognitive burden, we shortened the scale to 13 items. We retained the four items on self-efficacy and dropped the four items on critical consciousness, which were deemed to be more cognitively demanding. An example of an item of self-efficacy is, “You feel that you can change your daily schedule” (response options: completely disagree; partly disagree; partly agree; completely agree). For instrumental time-use agency, we followed the example of the Women’s Empowerment Metric for National Statistical Systems (WEMNS), in which the time-use agency module had been adapted by dropping items related to voice and revising the wording of the decision-making items to ask how much influence the respondent had over their time spent on nine different activities.[25, 26] An example of an item on decision-making is, “During the last 4 weeks, how much influence did you have in decisions about the amount of time you spent on Attending a community meeting?” (response options: no influence; a small amount of influence; a medium amount of influence; a lot of / high influence). The full modules for collection of data on time use and time-use agency are shown in Tables S1 and S2.

As noted above, the present study was nested within a larger study, and data collection took place within the context of the larger study activities, as shown in Figure 1. Data were collected in person from July 8 – August 16, 2024 in Kenya and July 22 – August 8, 2024 in Honduras. In Kenya, the data collection team consisted of 12 data collectors (eight women and four men) who were trained by the Emory team with support from St. Paul’s University. In Honduras, the data collection team consisted of 12 data collectors (eight women and four men) who were trained by the Emory team with support from the Edwin Roldan Medina Lopez Research Group. Data collectors were fluent in the local language (Samburu in Kenya and Spanish in Honduras) and were gender matched to participants. Data collection was done using paper forms, and all interviews were audio recorded for quality control. For women, because the research included data collection activities to assess women’s experiences collecting water, field team members worked in pairs, with one person collecting data and the other taking notes throughout the data collection process. For men, a single data collector audio recorded the interview, collected data, and took additional notes as needed. At the end of each interview, the data collector(s) reviewed the activity list together with the respondent for completeness. Audio recordings were translated and transcribed into English by the research teams in Kenya and Honduras, and transcripts were checked and validated.

**Fig 1.**
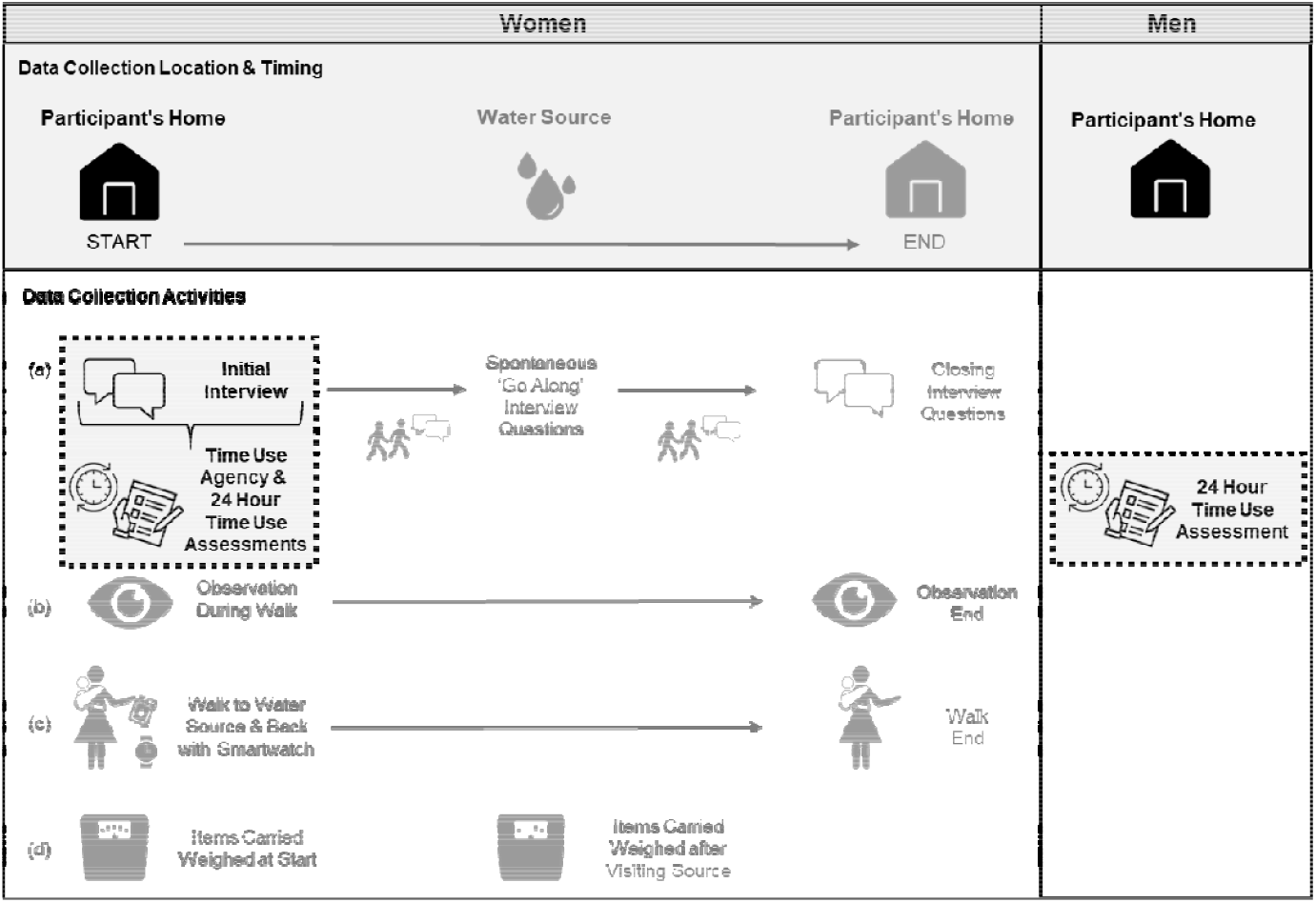
Data collection activities for the present study (shown in black) as nested within a larger study of women’s water collection experiences (activities shown in grey)

### Quality control and data management

Data from the paper forms were independently entered into an Excel spreadsheet by two different team members and reconciled to ensure data entry accuracy. In case of discrepancies, teams checked the audio recording. In reviewing data, the team identified issues with the translation of the time-use agency module in Honduras, in which the response option related to having ‘a lot of influence’ over one’s own time was translated / interpreted as ‘many people’ having influence over the participant’s time. Given that the meaning of this phrase was substantially changed, we decided to exclude the Honduras time-use agency data from further analysis.

Once data had been entered, they underwent another round of review and quality control. The time-use survey module included an option for “other” activities, in which data collectors could write open-ended responses. These open-ended responses were reviewed and, where possible, assigned to their appropriate activity categories. In some cases, the team went back to the transcript or audio recording to get additional context to inform the recategorization. As a further quality control step, we also reviewed a selection of transcripts against the quantitative time-use data to ensure that the data were aligned. Special attention was paid to examples of men’s water collection activities, particularly because it is known to be rare for men to engage in water collection, especially in Kenya. In Kenya, there were three men for whom ‘water fetching’ was recorded as an activity. We reviewed those transcripts to identify any additional context / description of their water collection activities. As a result, some instances were recategorized (digging a dam [unpaid work] and watering goats [livestock]). Due to these discrepancies, a decision was made to review all men’s transcripts from Kenya and to recategorize activities as needed. For men’s data from Honduras and women’s data from both Kenya and Honduras, we first restricted the dataset to those individuals who reported collecting water and then randomly selected one transcript from each enumerator team (5 in Kenya, 4 in Honduras) for review. This review identified very few discrepancies. Therefore, we determined that a full review of all transcripts was not needed.

As a next data management step, we aggregated and re-categorized the 27 activities from the activity list in our data collection tool, following published guidance from the UN Statistical Division. The UN Statistical Division’s Minimum Harmonized Instrument has 25 activity categories, which can be amended to conform to local practices.[27] We harmonized our categories where possible and created 17 activity categories, which we used for our primary analyses. We then further combined some activities to create seven larger categories (water collection, paid labor, unpaid labor, self-care, social activities, travel, and other), which we used for secondary analyses. Table 1 shows the three categorization schemes.

**Table 1.**
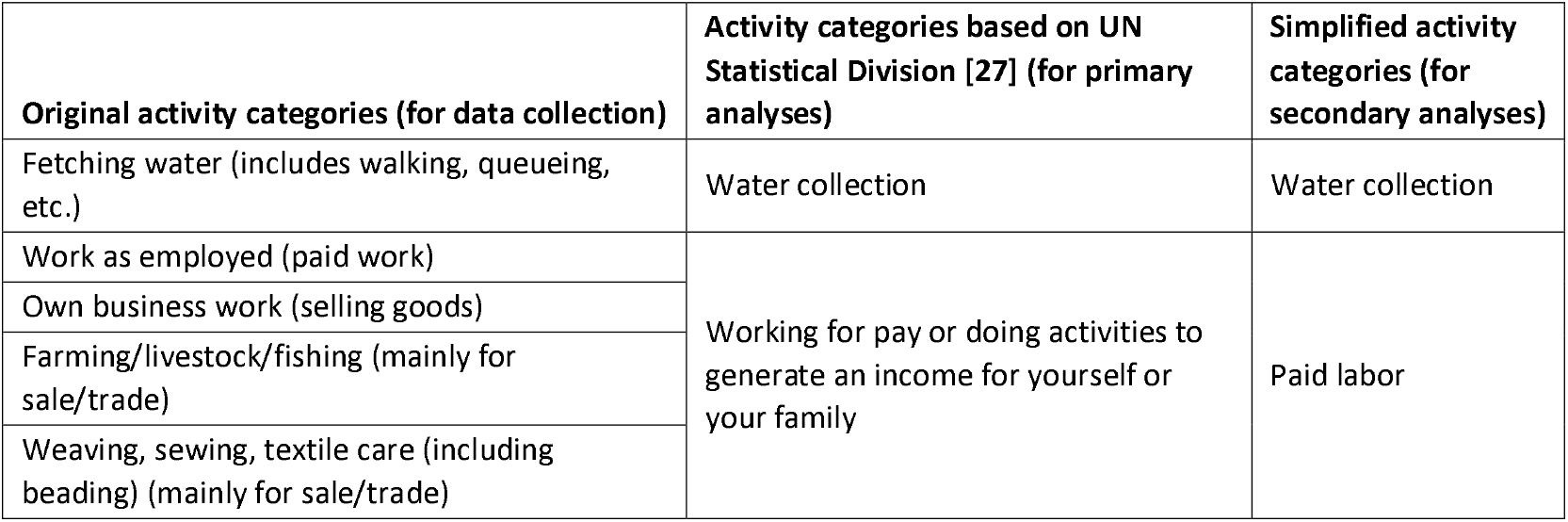
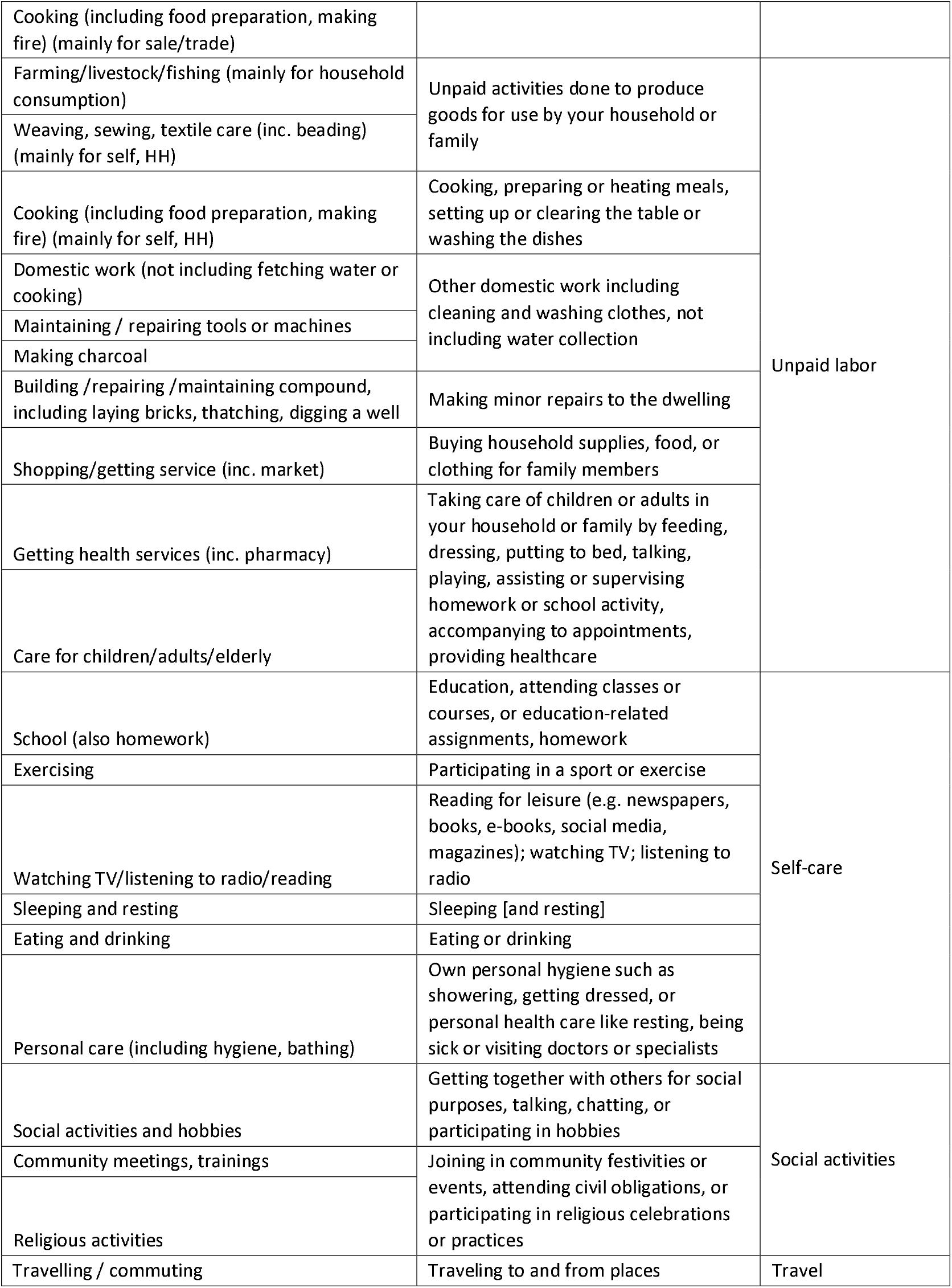
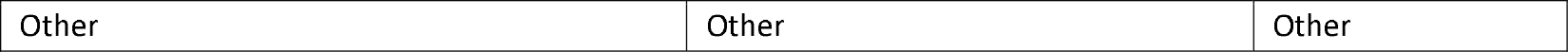
Categorization schemes of activities reported by participants used for data collection and analysis.

### Statistical analysis

We calculated descriptive statistics, including demographic characteristics of the study population and the total number of activities reported by women and men (excluding sleeping, which was reported by all participants), by study setting. Using the 17 activity categories based on the UN Statistical Division, we calculated the proportion of individuals who participated in each type of activity and the average (mean) time in minutes spent on each activity type, by gender and by type of water source, among those who participated in the activity and among the whole sample. We conducted chi-square tests to compare the proportion of individuals who participated in each type of activity by gender and by type of water source. We also conducted t-tests of mean time spent on each activity to compare differences by gender and by type of water source.

In secondary analyses of time use data, we used the seven larger activity categories and calculated mean time spent on each activity category by gender and type of water source. We also graphed the seven activity categories across the 24-hour data collection period for women and men, to examine broad patterns in women’s and men’s activities over the course of one day.

When measuring a construct that is latent (i.e., not directly observable), it is important to assess construct validity, or whether it is measuring what it is intended to measure.[28] Given that we aimed to measure the latent construct of time-use agency, we conducted a confirmatory factor analysis (CFA) to assess the construct validity of our time-use agency measure. We hypothesized a two-factor solution representing intrinsic and instrumental time-use agency, based on theory and empirical evidence from previous factor analyses. Published guidance suggests a sample size of about 5-10 subjects per item for factor analysis[29, 30], which would equate to a sample size of 65-130 respondents for our 13 items. Therefore, we determined that our sample size was adequate for a CFA. After running the CFA, we examined factor loadings and interpreted model fit based on the following indices: root mean squared error of approximation (RMSEA), comparative fit index (CFI), Tucker–Lewis index (TLI), and standardized root mean squared residual (SRMR). RMSEA < 0.08, CFI > 0.95, TLI > 0.95, and SRMR < 0.08 were considered good fit.[31] Due to the limited sample size, we did not run further analyses, such as multi-group confirmatory factor analysis, which would be necessary to validly compare scores between men and women.

The CFA was conducted in Stata version 19.0. All other analyses were conducted in R.

### Ethics

Ethical approvals were provided by the institutional review boards of Emory University (Atlanta, GA: STUDY00005955), Universidad Nacional Autónoma de Honduras (Tegucigalpa: CEIFCS-2023-P18), and St. Paul University (Nairobi: SPU/29/2023). A research license was granted by Kenya’s National Commission for Science, Technology, and Innovation (Nairobi: NACOSTI/P/24/3752). Research assistants read consent forms out loud to participants, who provided verbal consent, affirmed by signature of research assistant.

### Role of the funding source

The funder was involved in selecting study locations, including the study communities in each country. The funder had no other role in study design, data collection, data analysis, data interpretation, decision to publish, or preparation of the manuscript.

## RESULTS

### Participant characteristics

In Kenya, we collected data from 47 men and 48 women (95 total participants). Mean age for men was 45 years and for women was 37 years. All (100%) of the men were married, while among the women, 81% were married, 17% were widowed, and 2% were single. The majority of men (58%) and women (90%) had never attended school. The majority of both men and women reported an unimproved source (surface water or an unprotected dug well) as their primary source of water for drinking and other uses, and all (100%) said that their primary source was outside their dwelling or plot. On average, women performed more activities per day than men (6.5 women, 4.9 men), including more unpaid activities (2.7 women, 1.3 men). Additional details of participant characteristics are shown in Table 2.

**Table 2.**
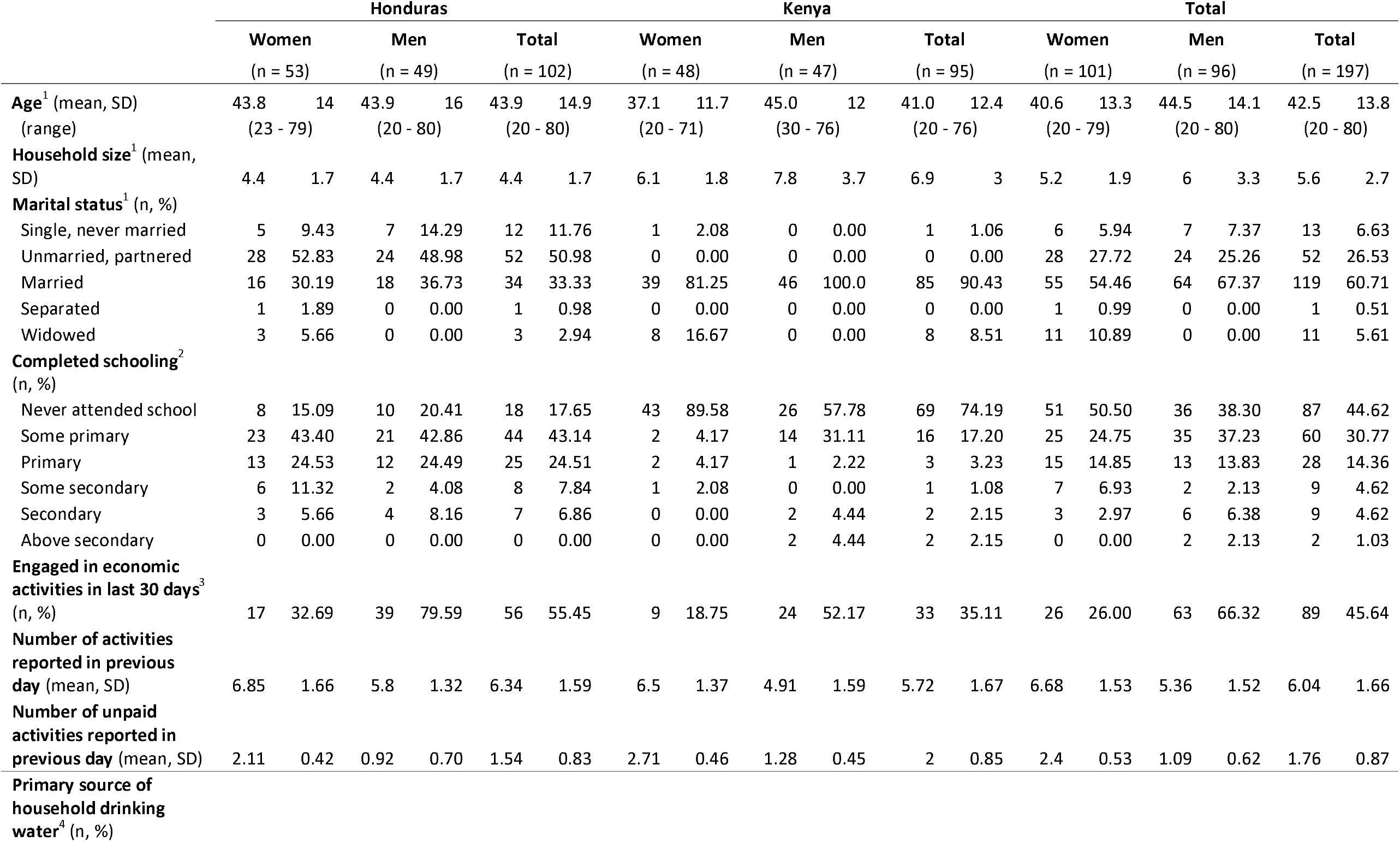
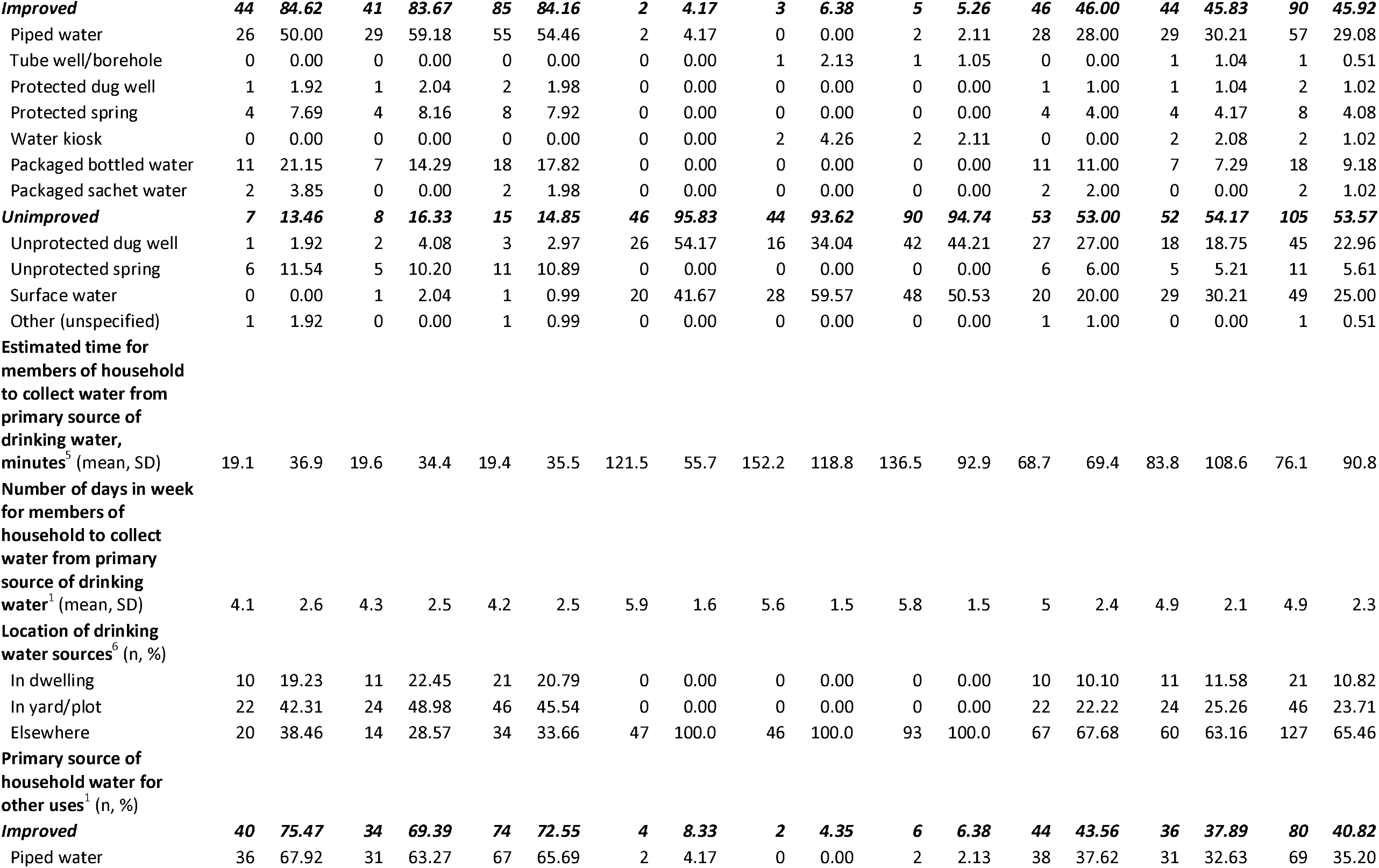
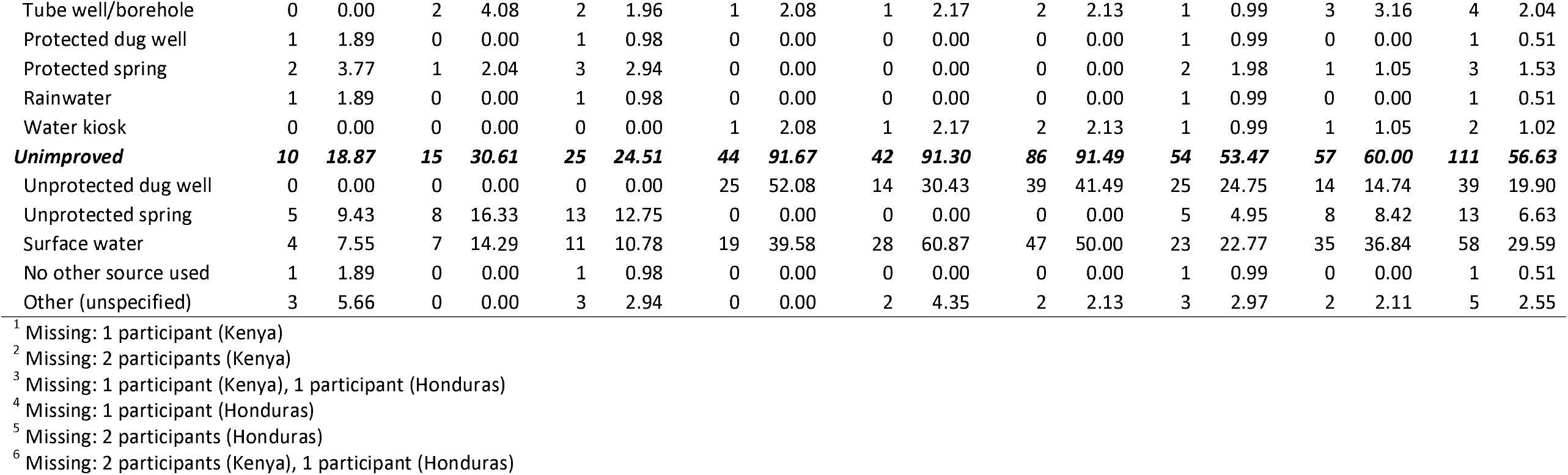
Participant characteristics, by country setting and by gender.

In Honduras, we collected data from 49 men and 53 women (102 total participants). Mean age for both men and women was 44 years. Among men and women, the largest proportion were unmarried but had a partner (49% men, 53% women). Most participants had some primary education or less (63% men, 58% women). Many participants reported an improved source (most often piped water or packaged bottled water) as their primary source of water for drinking and other uses, typically within their dwelling or plot. As in the Kenya sample, women performed more activities per day than men (6.9 women, 5.8 men), including more unpaid activities (2.1 women, 0.9 men) (Table 2).

### Time use by activity category, and proportion of time spent on each category

#### Kenya: Comparison of time use by gender

When comparing the proportion of individuals who participated in each type of activity, the chi-square tests indicated several significant differences between women and men (Table 3). Significantly larger proportions of women compared to men reported participating in water collection (68.8% (33/48) of women, 4.3% (2/47) of men, p<0.001); cooking, including food preparation and washing dishes (100% (48/48) of women, 2.1% 1/47) of men, p<0.001); other domestic work such as cleaning or laundry (100% (48/48) of women, 29.8% (14/47) of men, p<0.001); caregiving for children or adults in the household (31.5% (15/48) of women, 6.4% (3/47) of men, p=0.002); and “other” activities (35.4% (17/48) of women, 21.3% (6/47) of men, p=0.019). On the other hand, significantly larger proportions of men compared to women reported spending time on unpaid activities done to produce goods for use by their own household or family (70.8% (34/48) of women, 95.7% (45/47) of men; p=0.003); making minor repairs to the dwelling (2.1% (1/48) of women, 31.9% (15/47) of men, p<0.001); joining in community festivities or events, attending civil obligations, or participating in religious celebrations or practices (18.8% (9/48) of women, 42.5% (20/47) of men, p=0.022); and traveling (16.7% (8/48) of women, 63.8% (30/48) of men, p<0.001).

**Table 3.** Time use by activity category, and proportion of time spent on each category, for women and men in Kenya sample.

| Activity | Women (N=48) |  |  | Men (N=47) |  |  |  |  |
| --- | --- | --- | --- | --- | --- | --- | --- | --- |
|  | Number who participated | % of women who participated | Average hours spent on activity<sup>1</sup> | Number who participated | % of men who participated | Average hours spent on activity<sup>1</sup> | p-value for chi-square test<sup>2</sup> | p-value for t-test<sup>3</sup> |
| Working for pay or doing activities to generate an income for yourself or your family | 10 | 20.8 | 2.03 | 9 | 19.1 | 3.78 | 1.000 | 0.453 |
| Unpaid activities done to produce goods for use by your household or family | 34 | 70.8 | 5.65 | 45 | 95.7 | 7.13 | 0.003 | <0.001 |
| Water collection | 33 | 68.8 | 1.51 | 2 | 4.3 | 2.75 | <0.001 | <0.001 |
| Cooking, preparing or heating meals, setting up or clearing the table or washing the dishes | 48 | 100 | 2.90 | 1 | 2.1 | 1.00 | <0.001 | <0.001 |
| Other domestic work including cleaning and washing clothes, not including water collection | 48 | 100 | 5.64 | 14 | 29.8 | 2.45 | <0.001 | <0.001 |
| Making minor repairs to the dwelling | 1 | 2.1 | 2.50 | 15 | 31.9 | 2.07 | <0.001 | 0.001 |
| Buying household supplies, food, or clothing for family members | 3 | 5.3 | 2.42 | 6 | 12.8 | 1.42 | 0.317 | - |
| Taking care of children or adults in your household or family | 15 | 31.5 | 2.07 | 3 | 6.4 | 1.00 | 0.002 | 0.024 |
| Education: attending classes or courses or education related assignments/homework | 4 | 8.3 | 2.00 | 0 | 0 | 0.00 | 0.117 | - |
| Getting together with others for social purposes | 17 | 35.4 | 1.64 | 19 | 40.4 | 1.93 | 0.771 | 0.395 |
| Joining in community festivities or events | 9 | 18.8 | 2.39 | 20 | 42.6 | 3.58 | 0.022 | 0.008 |
| Participating in a sport or exercise | 1 | 2.1 | 0.75 | 2 | 4.3 | 2.25 | 0.617 | 0.252 |
| Reading for leisure, watching TV, listening to radio | 1 | 2.1 | 1.00 | 2 | 4.3 | 1.50 | 0.617 | - |
| Sleeping and resting | 48 | 100 | 7.27 | 47 | 100 | 9.13 | 1.000 | <0.001 |
| Eating or drinking | 46 | 95.8 | 1.53 | 46 | 97.9 | 1.75 | 1.000 | 0.271 |
| Own personal hygiene | 17 | 35.4 | 0.96 | 11 | 23.4 | 0.90 | 0.194 | 0.195 |
| Travel to and from places | 8 | 16.7 | 1.88 | 30 | 63.8 | 2.32 | <0.001 | <0.001 |
| Other activities* | 17 | 35.4 | 1.22 | 6 | 12.8 | 1.21 | 0.019 | 0.046 |

<sup>1</sup> Average hours are calculated as the mean hours spent among those who reported spending any (non-zero) amount of time on the activity
<sup>2</sup> Chi-square test to compare the proportions of individuals who participated in each activity by gender
<sup>3</sup> T-test to compare mean time spent on activity by gender among all participants

Concurrently, among this sample of men and women from Kenya, we observed that women reported spending significantly more time, on average, than men on water collection (1.04 hours for women, 0.12 hours for men, p<0.001); cooking (2.90 hours for women, 0.02 hours for men, p<0.001); other domestic work such as cleaning or laundry (5.64 hours for women, 0.73 hours for men, p<0.001); caregiving for children or adults in the household (0.65 hours for women, 0.04 hours for men, p=0.024); and “other” activities (0.43 hours for women, 0.15 hours for men, p=0.046) (Figure 2). Men’s self-reported average time spent was significantly higher than women’s for unpaid activities done to produce goods for use by their own household or family (4.01 hours for women, 6.82 hours for men, p<0.001); making minor repairs to the dwelling (0.05 hours for women, 0.66 hours for men, p=0.001); joining in community festivities or events, attending civil obligations, or participating in religious celebrations or practices (0.45 hours for women, 1.53 hours for men, p=0.008); traveling (0.31 hours for women, 1.48 hours for men, p<0.001); and sleeping (7.27 hours for women, 9.13 hours for men, p<0.001) (Figure 2).

**Fig 2.**
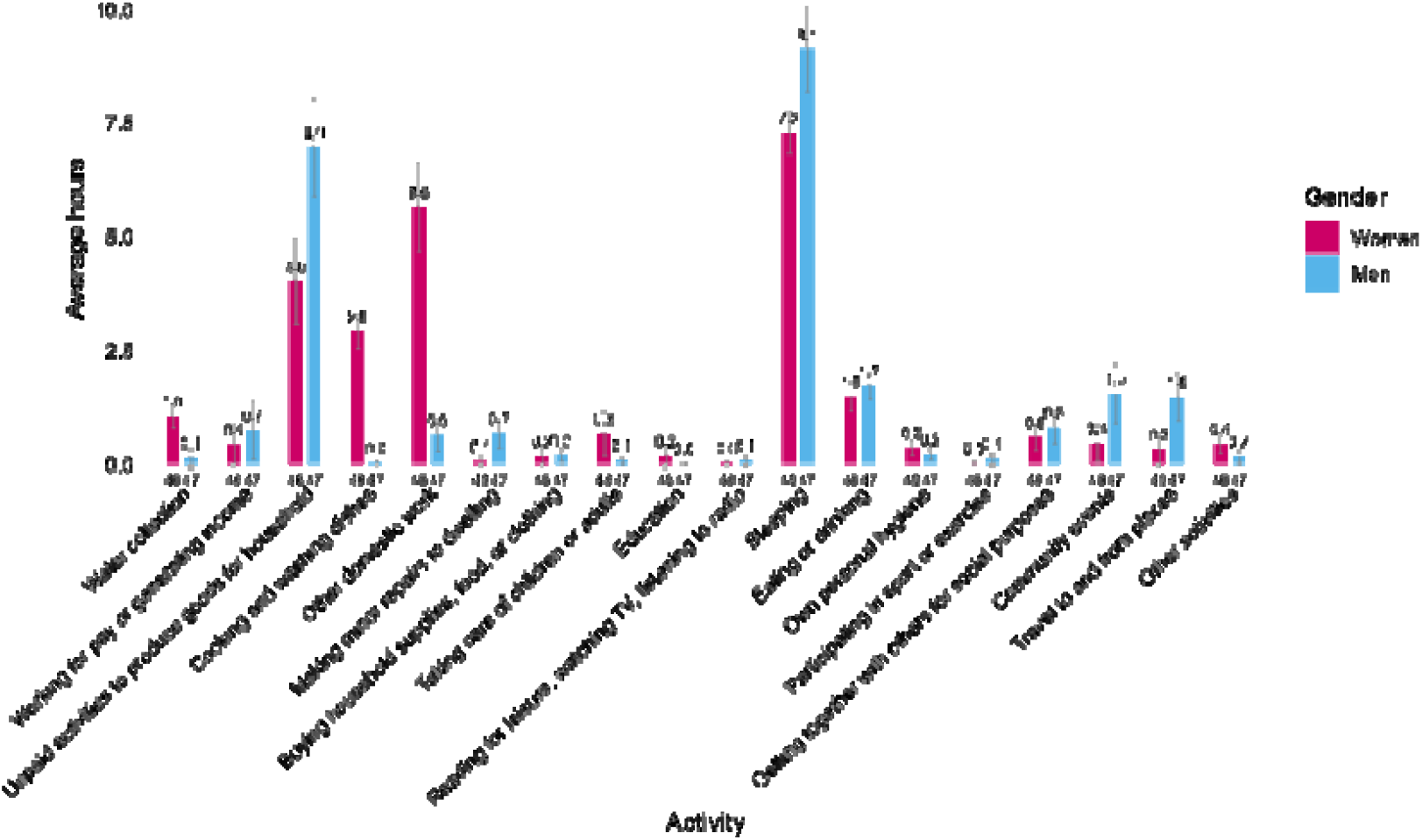
Average self-reported time use in hours among all participants in Kenya sample (N=95), by activity type and gender ***Note:*** Numbers below bars indicate the number of respondents; numbers above bars indicate the average time (hours) spent on the activity

When restricting the sample to only those who participated in each activity, fewer differences emerged. Women still spent significantly more time than men on other domestic work such as cleaning or laundry (p<0.001) and also spent more time on unpaid activities done to produce goods for use by their own household or family (p=0.045), whereas men still spent significantly more time than women sleeping (p<0.001) (S1 Figure).

In secondary analyses using the seven larger activity categories, results from the comparison of women’s and men’s time allocation were similar to those from the primary analyses, with women spending significantly more time than men on water collection and unpaid labor and men spending significantly more time than women on selfcare, social activities, and travel (S2 Figure). Graphs of the seven activity categories across the 24-hour data collection period for women and men showed that unpaid labor was the dominant activity throughout the day for women, with participation peaking in the early morning (hours 5–7) and again in the late afternoon and evening (hours 18–19) (S3 Figure).

#### Kenya: Comparison of women’s time use by intervention status

Among women, when comparing the proportion who reported participating in each activity by intervention status of the community, we observed only one significant difference: a larger proportion of women in intervention communities (which had received improved water sources through World Vision) reported joining in community festivities or events, attending civil obligations, or participating in religious celebrations or practices, compared with women in comparison communities (which had not yet received improved water sources from World Vision) (33.3% in intervention, 4.3% in comparison, p=0.027) (Table 4). On the other hand, women in comparison communities reported spending significantly more time getting together with others for social purposes, talking, chatting, or participating in hobbies, compared with women in intervention communities (0.93 hours in intervention, 2.12 hours in comparison, p=0.031). We did not observe any other significant difference in activities or time allocation, including time spent on water collection, on working for pay or income-generating activities, or on any other form of unpaid work, between women in each type of community.

**Table 4.**
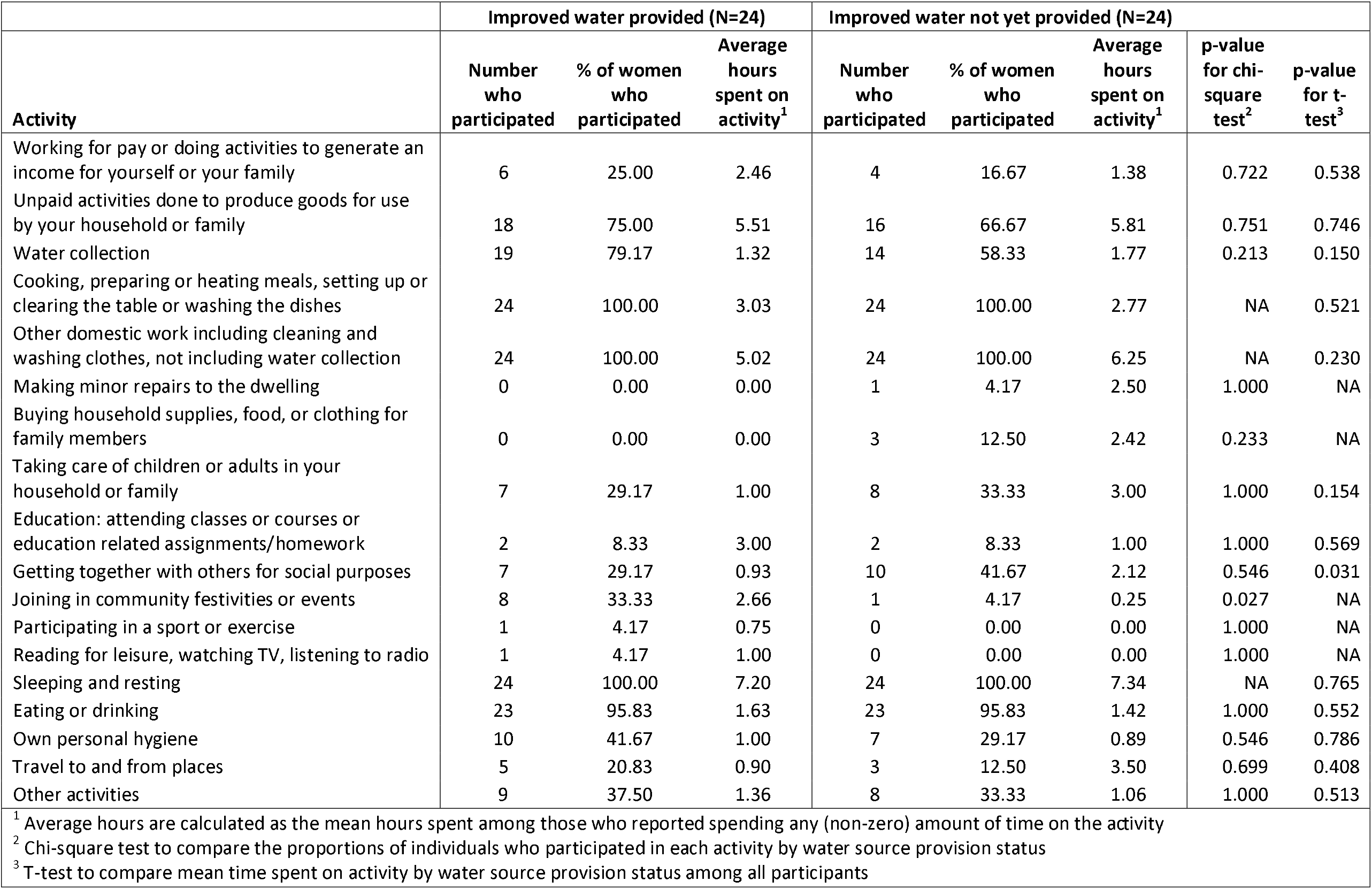
Time use by activity category, and proportion of time spent on each category, for women by community type in Kenya sample.

In the secondary analysis of women’s time allocation between intervention and comparison communities using the seven larger activity categories, no significant differences emerged (S4 Figure).

#### Honduras: Comparison of time use by gender

When comparing the proportion of individuals who participated in each type of activity, the chi-square tests indicated significant differences between women and men for 9 of 16 activity categories in which any individuals reported participating (Table 5). Significantly larger proportions of women compared to men reported participating in cooking, including food preparation and washing dishes (100% (53/53) of women, 2.0% (1/49) of men, p<0.001); other domestic work such as cleaning or laundry (96.2% (51/53) of women, 53.1% (26/49) of men, p<0.001); caregiving for children or adults in the household (49.1% (26/53) of women, 10.2% (5/49)of men, p<0.001); joining in community festivities or events, attending civil obligations, or participating in religious celebrations or practices (50.9% (27/53) of women, 26.5% (13/49) of men, p=0.020); and “other” activities (15.1% (8/53) of women, 2.0% (1/49) of men, p=0.049) On the other hand, significantly larger proportions of men compared to women reported spending time working for pay or generating an income (18.9% (10/53)of women, 65.3% (13/49) of men; p=<0.001); doing unpaid activities to produce goods for use by their own household or family (15.1% (8/53) of women, 36.7% (18/49) of men; p=0.023); eating or drinking (86.8% (46/53) of women, 100% (49/49 of men; p=0.025); and traveling (60.4% (32/53) of women, 85.7% (42/49) of men, p=0.008). We did not observe any differences in time spent on water collection between men and women (9.4% (5/53) of women, 4.1% (2/49) of men, p=0.499).

**Table 5.** Time use by activity category, and proportion of time spent on each category, for women and men in Honduras sample.

| Activity | Women (N=53) |  |  | Men (N=49) |  |  |  |  |
| --- | --- | --- | --- | --- | --- | --- | --- | --- |
|  | Number who participated | % of women who participated | Average hours spent on activity<sup>1</sup> | Number who participated | % of men who participated | Average hours spent on activity<sup>1</sup> | p-value for chi-square test<sup>2</sup> | p-value for t-test<sup>3</sup> |
| Working for pay or doing activities to generate an income for yourself or your family | 10 | 18.87 | 2.80 | 32 | 65.31 | 6.60 | <0.001 | <0.001 |
| Unpaid activities done to produce goods for use by your household or family | 8 | 15.09 | 0.56 | 18 | 36.73 | 2.29 | 0.023 | 0.002 |
| Water collection | 5 | 9.43 | 0.65 | 2 | 4.08 | 0.38 | 0.499 | 0.147 |
| Cooking, preparing or heating meals, setting up or clearing the table or washing the dishes | 53 | 100 | 2.79 | 1 | 2.04 | 0.25 | <0.001 | <0.001 |
| Other domestic work including cleaning and washing clothes, not including water collection | 51 | 96.23 | 2.86 | 26 | 53.06 | 1.38 | <0.001 | <0.001 |
| Making minor repairs to the dwelling | NA | NA | NA | 2 | 4.08 | 3.62 | 0.441 | NA |
| Buying household supplies, food, or clothing for family members | 6 | 11.32 | 0.67 | 5 | 10.20 | 2.20 | 1.000 | 0.223 |
| Taking care of children or adults in your household or family | 26 | 49.06 | 2.14 | 5 | 10.20 | 1.80 | <0.001 | 0.001 |
| Education: attending classes or courses or education related assignments/homework | NA | NA | NA | NA | NA | NA | NA | NA |
| Getting together with others for social purposes | 31 | 58.49 | 2.15 | 31 | 63.27 | 2.85 | 0.771 | 0.155 |
| Joining in community festivities or events | 27 | 50.94 | 1.91 | 13 | 26.53 | 2.79 | 0.020 | 0.448 |
| Participating in a sport or exercise | NA | NA | NA | NA | NA | NA | NA | NA |
| Reading for leisure, watching TV, listening to radio | 13 | 24.53 | 1.56 | 20 | 40.82 | 2.31 | 0.122 | 0.037 |
| Sleeping and resting | 53 | 100 | 11.15 | 48 | 97.96 | 10.69 | 0.969 | 0.341 |
| Eating or drinking | 46 | 86.79 | 1.12 | 49 | 100.00 | 1.06 | 0.025 | 0.569 |
| Own personal hygiene | 47 | 88.68 | 0.78 | 37 | 75.51 | 0.63 | 0.138 | 0.026 |
| Travel to and from places | 32 | 60.38 | 1.23 | 42 | 85.71 | 2.31 | 0.008 | <0.001 |
| Other activities | 8 | 15.09 | 3.31 | 1 | 2.04 | 3.00 | 0.049 | 0.093 |

<sup>1</sup> Average hours are calculated as the mean hours spent among those who reported spending any (non-zero) amount of time on the activity <sup>2</sup> Chi-square test to compare the proportions of individuals who participated in each activity by gender <sup>3</sup> T-test to compare mean time spent on activity by gender among all participants

Concomitantly, among this sample of men and women from Honduras, we observed that women reported spending significantly more time, on average, than men on cooking (2.79 hours for women, 0.01 hours for men, p<0.001); other domestic work such as cleaning or laundry (2.75 hours for women, 0.73 hours for men, p<0.001); caregiving for children or adults in the household (1.05 hours for women, 0.18 hours for men, p=0.024); and their own personal hygiene (0.69 hours for women, 0.47 hours for men, p=0.026) (Figure 3). Men’s self-reported average time spent was significantly higher than women’s on working for pay or generating income (0.53 hours for women, 4.31 hours for men, p<0.001); unpaid activities done to produce goods for use by their own household or family (0.08 hours for women, 0.84 hours for men, p=0.002); reading for leisure, watching TV, or listening to the radio (0.38 hours for women, 0.94 hours for men; p=0.037); and traveling (0.75 hours for women, 1.98 hours for men, p<0.001) (Figure 3).

**Fig 3.**
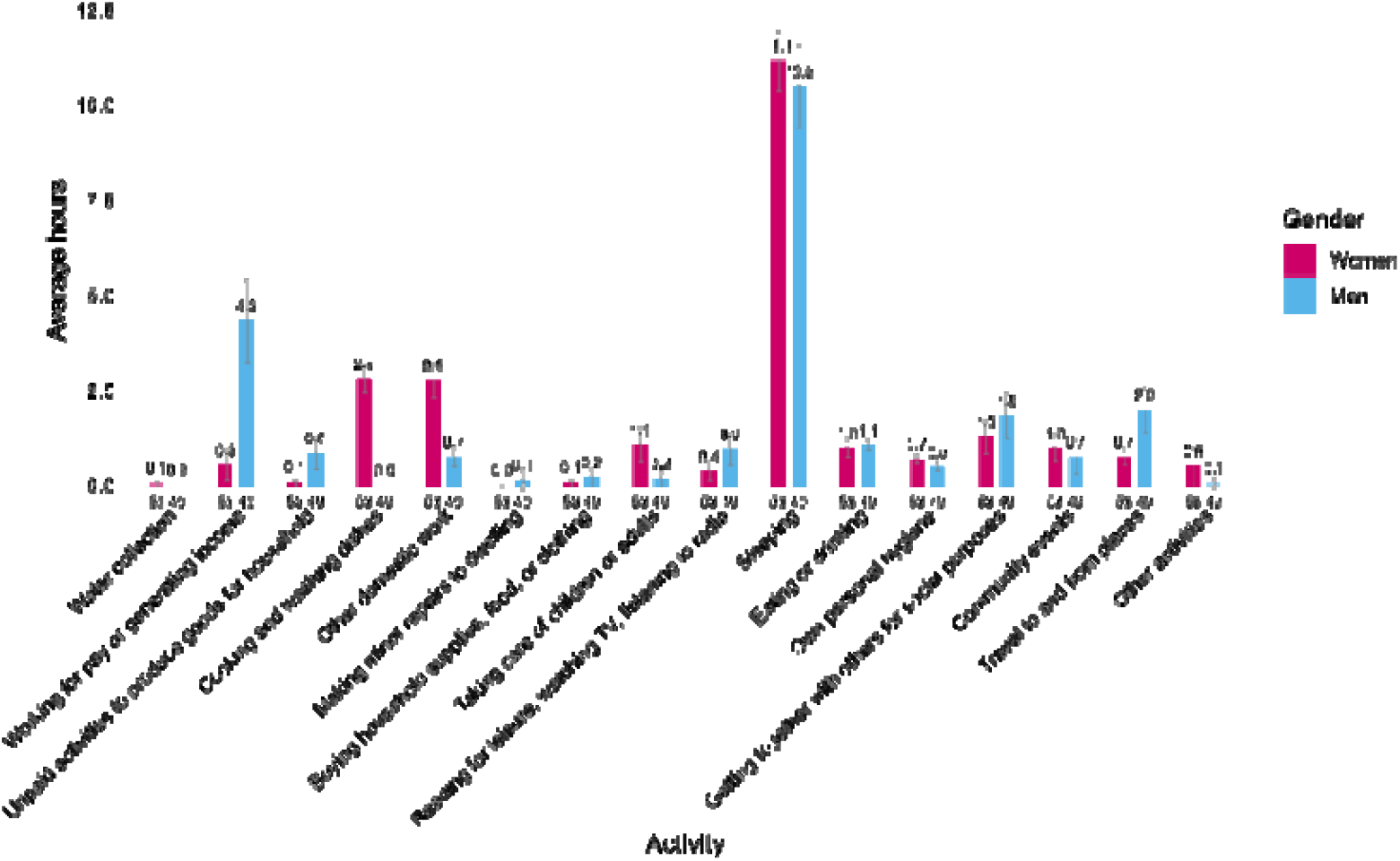
Average self-reported time use in hours among all participants in Honduras sample (N=102), by activity type and gender ***Note:*** Numbers below bars indicate the number of respondents; numbers above bars indicate the average time spent on the activity

When restricting the sample to only those who participated in each activity, women still spent significantly more time than men on other domestic work such as cleaning or laundry (p<0.001), whereas men still spent significantly more time than women working for pay or generating income (p=0.001); doing unpaid activities to produce goods for use by their own household or family (p=0.002); and traveling to and from places (p=0.005) (S5 Figure).

In secondary analyses using the seven larger activity categories, results from the comparison of women’s and men’s time allocation indicated that women spent significantly more time than men on unpaid labor, while men spent significantly more time than women on paid labor; no other differences were observed between genders (S6 Figure). Graphs of the seven activity categories across the 24-hour data collection period for women and men showed that in Honduras, men had relatively high participation in paid labor from hours 6 through 12, peaking mid-morning. For women, unpaid labor dominated the day with a peak around hours 7–8 and high participation through midday, followed by a secondary rise in social activities in the afternoon and evening hours. Social activities were notably more prevalent in Honduras than in Kenya for both men and women, particularly in the afternoon, while water collection was minimal (S7 Figure).

#### Honduras: Comparison of women’s time use by intervention status

Among women, when comparing the proportion who reported participating in each activity by intervention status of the community, we observed three significant differences. First, none of the women in intervention communities reported doing unpaid activities to produce goods for use by their own household or family, while almost one-quarter of women in comparison communities did so (0% in intervention, 23.5% in comparison, p=0.040). Second and third, a larger proportion of women in intervention communities reported getting together with others for social purposes, talking, chatting, or participating in hobbies (79.0% in intervention, 47.1% in comparison; p=0.049) and reading for leisure, watching TV, or listening to the radio (47.4% in intervention, 11.8% in comparison, p=0.011) (Table 6). In comparing time spent on each activity, we observed that women in intervention communities spent significantly more time getting together with others for social purposes, talking, chatting, or participating in hobbies (2.73 hours in intervention, 1.59 hours in comparison, p=0.006) and significantly less time sleeping (10.12 hours in intervention, 11.72 hours in comparison, p=0.037) than women in comparison communities. We did not observe any other significant difference in activities or time allocation, including time spent on water collection, on working for pay or income-generating activities, or on any other form of unpaid work, between women in each type of community.

**Table 6.**
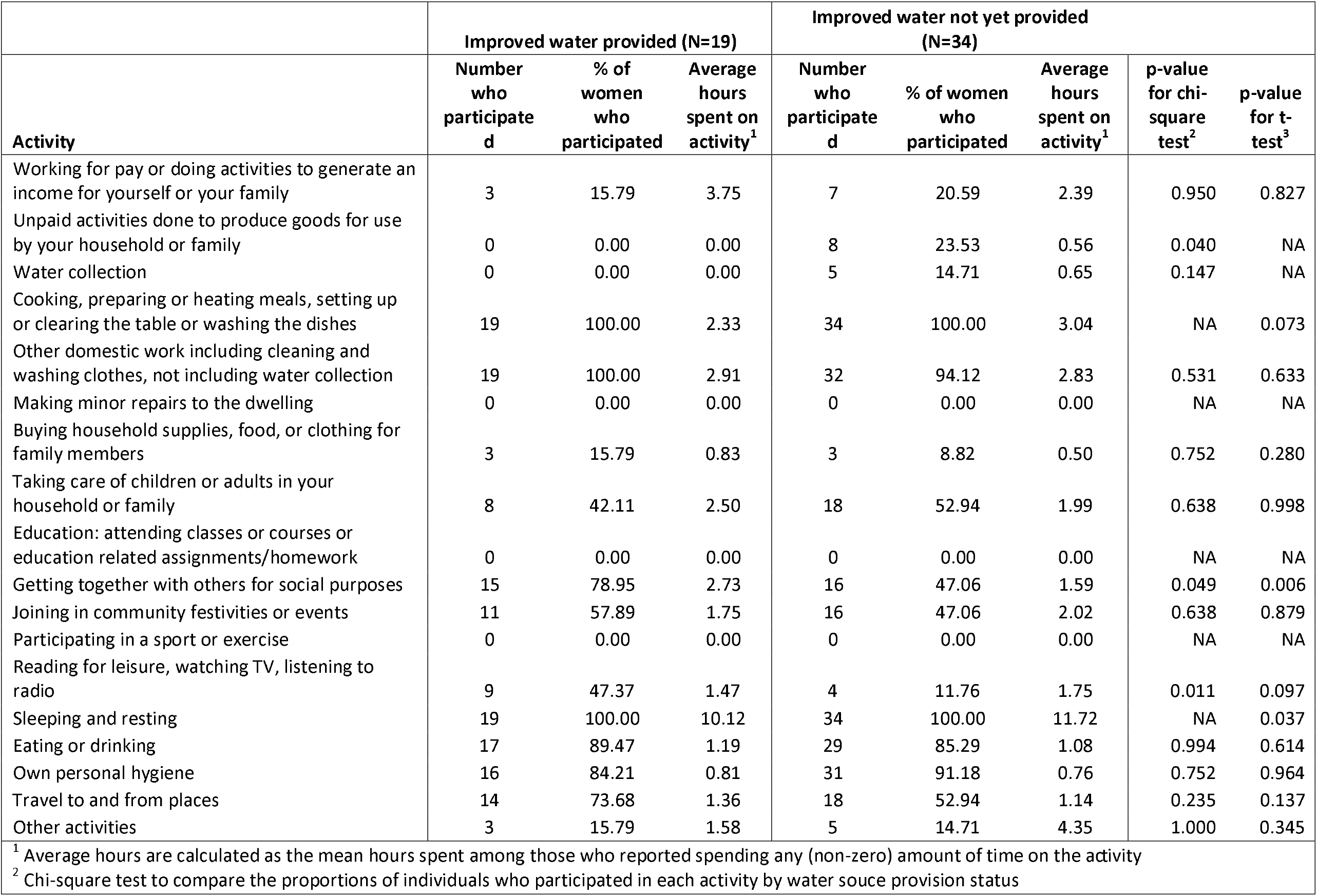
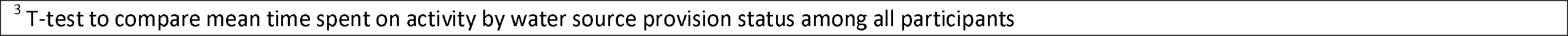
Time use by activity category, and proportion of time spent on each category, for women by community type in Honduras sample.

In the secondary analysis of women’s time allocation between intervention and comparison communities using the seven larger activity categories, no significant differences emerged (S8 Figure).

### Time-use agency

As noted above, results for time-use agency are restricted to data from Kenya. Among items designed to measure intrinsic time-use agency (defined for our purposes as self-efficacy), more than three-quarters of men and women partly or completely agreed with all four items (Table S3). The largest discrepancy between men and women was in their perceived ability to change the amount of time they spend on paid work, with men being more likely to ‘completely disagree’ (Table S3). Among items that asked about the amount of influence the respondent had over the amount of time they spent on different activities, a majority of respondents said that they had a high degree of influence over their time spent on all listed activities. The only exception was the item that asked about time spent on household duties, for which less than half (43.5%; 20/46) of men said that they had a high degree of influence (women: 97.9% (46/47) (Table S3).

In the initial two-factor CFA, three of the four items for the first factor (TU1, TU2, and TU4, which were designed to measure aspects of intrinsic time-use agency) had low loadings (<|0.300|), indicating that they had little shared variance (i.e., a weak relationship with each other and any underlying construct). We dropped items one at a time and iteratively re-ran the CFA with the remaining items. In the end, given that it is not possible to have a factor with only one item, we dropped all items from the first factor and re-ran the CFA with only the second factor, which was designed to measure instrumental time-use agency. We dropped items based on low loadings (TU5, TU6, and TU12) and iteratively re-ran the CFA with the remaining items until a satisfactory factor structure was achieved. The final factor solution has six items and excellent model fit (Table 7).

**Table 7.**
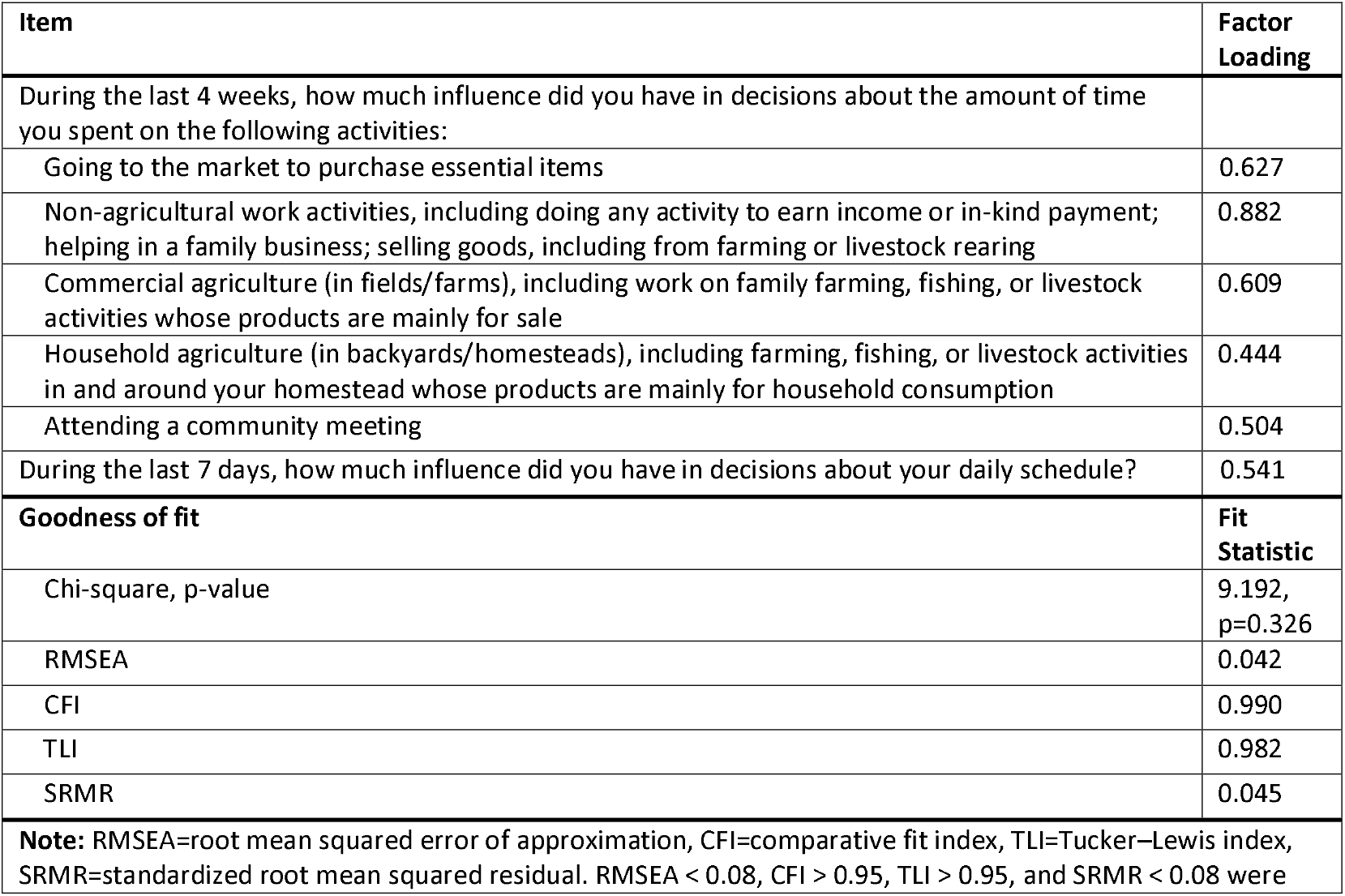
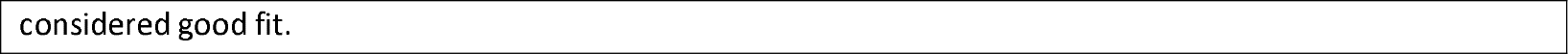
Results from confirmatory factor analysis of items representing instrumental time-use agency.

| Item | Factor Loading |
| --- | --- |
| During the last 4 weeks, how much influence did you have in decisions about the amount of time you spent on the following activities: |  |
| Going to the market to purchase essential items | 0.627 |
| Non-agricultural work activities, including doing any activity to earn income or in-kind payment; helping in a family business; selling goods, including from farming or livestock rearing | 0.882 |
| Commercial agriculture (in fields/farms), including work on family farming, fishing, or livestock activities whose products are mainly for sale | 0.609 |
| Household agriculture (in backyards/homesteads), including farming, fishing, or livestock activities in and around your homestead whose products are mainly for household consumption | 0.444 |
| Attending a community meeting | 0.504 |
| During the last 7 days, how much influence did you have in decisions about your daily schedule? | 0.541 |
| **Goodness of fit** | **Fit Statistic** |
| Chi-square, p-value | 9.192, p=0.326 |
| RMSEA | 0.042 |
| CFI | 0.990 |
| TLI | 0.982 |
| SRMR | 0.045 |
| **Note:** RMSEA=root mean squared error of approximation, CFI=comparative fit index, TLI=Tucker–Lewis index, SRMR=standardized root mean squared residual. RMSEA < 0.08, CFI > 0.95, TLI > 0.95, and SRMR < 0.08 were |  |

considered good fit.

## DISCUSSION

In our study populations in rural Kenya and Honduras, we observed significant differences in time use between women and men, with women spending significantly larger amounts of time than men on unpaid domestic activities and care work such as cooking, cleaning, laundry, and caregiving for children or other adults in the household. In Kenya, we additionally observed that women spent significantly more time on water collection than men, while men spent significantly more time sleeping than women. In Kenya, there was no significant difference in the proportion of men and women who reported working for pay, while in Honduras, men were significantly more likely than women to have done so. When comparing women in communities that had or had not yet received improved water sources through a World Vision intervention, we observed no significant differences in either Kenya or Honduras in women’s time spent on water collection or on income-generating activities. With respect to time-use agency, both women and men in Kenya reported having high degrees of influence over their time. Finally, we identified a concise set of six items to measure instrumental time-use agency among women and men in Kenya, which can be tested further in future research.

Overall, the data on differences in time allocation between women and men are closely aligned with data on time use from around the world. Kenya is among the few countries for which nationally representative data are available, as the Kenya National Bureau of Statistics (KNBS) undertook its first-ever national time use survey in 2021. Results of that survey show that the proportion of time spent on unpaid care work per day is approximately seven times higher for women (2.4%) than men (0.4%).[32] Similarly, the proportion of time spent on unpaid domestic work is about five times higher for women (16.3%) than for men (3.2%)[32]. While the KNBS data and our data are not directly comparable, our results similarly found that the number of hours spent per day caring for children and others in the household was six times higher for women (0.6 hours) than for men (0.1 hour) and the number of hours spent on water collection, cooking, and other unpaid domestic work was almost 14 times higher for women (9.5 hours) than for men (0.7 hours). Similar nationally representative data are lacking for Honduras, but our data on water collection are also broadly similar to global data, indicating that women and girls bear primary responsibility for this task in most settings.[20]

It is surprising that we did not observe any differences between women in intervention *versus* comparison communities in time spent on water collection or income-generating activities. This finding does not support the theory of change underlying World Vision’s SWSW program, which posits that provision of improved water services will lead to time savings for women and enable their involvement in economic activities. The theory of change was based on previous evidence, including from another World Vision project in Zambia, in which a piped water intervention led to time savings for women and increased time working outside the home.[33] The intervention in Zambia involved 20-25 taps being installed per village, with each tap being accessed by 2-5 households.[33] In our study, the intervention in Kenya was a community-level water kiosk and in Honduras was household-level piped water. However, in both Kenya and Honduras, we observed that women continued to use sources other than those provided by World Vision to meet their household needs.[21] Therefore, our results may be more similar to those of a study in rural Benin: when village-level water infrastructure was constructed that reduced distances to water sources, some households continued to use a more distant water source to avoid fees associated with the newer (closer) source.[34] Among those who used the newer source, some households collected more water per day due to the increased accessibility of the water source, and women still had to wait at the newer source, reducing their potential time savings.[34] Further, among our study population in Kenya, data were collected when there were unseasonal rains, which enabled women to collect unimproved surface water from sources that were closer to their homes than other options, including the improved sources from World Vision.[21] Ultimately, at the point in time when we collected data, women with the intervention spent the same amount of time collecting water as those without, leaving their time available for income-generating activities unchanged.

With respect to time-use agency, most women and men reported having a high degree of influence over their time spent on all listed activities. This result is promising, as it suggests that women’s agency is not the primary constraint on their time allocation. At the same time, while the CFA results demonstrated good fit for measurement of instrumental time-use agency, they also indicated that items intended to measure intrinsic time-use agency had poor construct validity, meaning that they did not appear to be measuring a salient latent construct. This was surprising, given that the same four items performed well when administered as part of a survey in Ghana.[24] It is possible that the construct of self-efficacy related to time use may be contextually sensitive.[35] Our findings reinforce the importance of ongoing validation when using items in new settings.[28] Future research may examine possible cross-cultural differences that affected the performance of these items in our Kenya sample as well as the utility of these items to measure time-use agency in other populations and contexts.

### Implications and recommendations for future programs, research, and policy

Our results suggest that program designers should not assume that provision of improved water supply, especially at the community level, will decrease women’s time spent on water collection. Rather, more transformative solutions may be needed that are deliberately designed to impact outcomes specific to women, including their time and labor. This recommendation is in line with transformative water, sanitation, and hygiene (WaSH) approaches, which promote infrastructure that provides continuous and convenient access to water for drinking and other household needs.[36] Ideally, as has been argued decades ago, piped connections should be provided over public resources.[37] Water supply should also be consistent and sufficient to meet all household needs, as research has shown that even with piped water, disruptions to supply lead women to reduce their paid working hours to wait at home for water.[38] Such transformative WaSH solutions would additionally contribute to SDG Target 6.1, “achieve universal and equitable access to safe and affordable drinking water for all”. Beyond infrastructure, our results further suggest a need for programs that aim to transform gendered social norms that assign water work almost exclusively to women and in which women’s time may be viewed as a collective--and free or low-cost--resource.[39, 40]

With respect to research, our results highlight the importance of measuring gender-specific outcomes, including but not limited to time use, from water interventions. It should be noted that this recommendation is not new; similar recommendations have been made for decades.[41-43] For example, in 2004, Roy and Crow wrote, “Future research will need to examine rural areas to determine precisely who collects water, how much time water collection consumes, and the quality of water available to each user. By gathering data on water collection and other work time of people in rural areas, researchers could discover the effect of water collection on how people allocate their time.”[42] Further, research could seek to advance assessment of water-related time burden beyond collection alone. A growing body of research has been investigating the time burden of water treatment, visiting water sources to carry out water-related work (e.g., laundry, washing cooking utensils), and the cognitive labor of household water management that also consumes women’s time.[7, 9, 44] Beyond the effects of water interventions on gender-specific time allocation, research should also measure effects on women’s health outcomes, including mental health and wellbeing, and on gendered social norms.[41] Further research is also needed on the measurement of time-use agency, including self-efficacy and other aspects of intrinsic time-use agency.

In terms of policy implications, our results reinforce the importance of collecting detailed data on time use for informing policy. Gender data on women’s time use and specifically on women’s unpaid care and domestic work remains sparse across global settings. As has been noted recently, quantifying unpaid and care work “is essential for designing gender-responsive policies and advancing inclusive development for all”.[4] When collected and used well, data on time use can inform policies and investments, including for infrastructure, services, and programs to reduce inequality and improve women’s lives.[4] In addition to producing more data, it is important to promote data use and communication among a broad range of stakeholders, including government, NGOs, and development partners.[45] Involvement of stakeholders beyond government could contribute to increasing public awareness and discourse about inequalities in time distribution within society as well as accountability towards social justice.[4]

### Strengths and limitations

Important strengths of our study include that we collected data in two low-resource contexts (Kenya and Honduras), in intervention and comparison communities, and among women and men. We collected data across a comprehensive range of activities, adapting the WEAI survey tool to measure water collection as a separate activity. At the same time, a common limitation of time use studies, including ours, is that data are collected for one 24-hour period, during which activities may not be representative of usual activities. We also collected data only during one season, meaning that we cannot draw conclusions about time use in other seasons or other climatic conditions. As noted above, in Kenya, the study site was experiencing unseasonal heavy rainfall at the time of data collection. Under typical arid conditions, women in these communities rely primarily on distant water sources.[7] However, during the study period, the availability of nearby unimproved water sources, made accessible by the unusual rainfall, may have reduced both the time required for water collection and the observed differences between intervention and comparison communities. Future research could aim to collect data over more days, across seasons, to better capture the variability in women’s time allocation related to water access and could include observation to reduce measurement error.

We also encountered several challenges related to data collection, including difficulties with measuring simultaneous activities, which are known to be systematically underestimated in time use studies.[46] Given that women in our study settings may typically combine unpaid and informal work with housework and childcare activities, the fact that we did not capture these simultaneous activities may have led to underestimation of women’s domestic and care work hours.[47] We observed that seven women in Honduras did not have any reported time spent eating or drinking in the previous day; based on debriefing sessions in the field, this was likely because women ate as a simultaneous and secondary activity while cooking or completing another task, meaning that eating was not captured in our data. In Kenya, another challenge involved the fact that our measurement of time use was based on clock time, which is a Western social construct that may not be locally appropriate.[48] Kenyan respondents’ conceptualization of time appeared more aligned with epochal time, which is based on events rather than clocks, requiring data collectors to ‘translate’ responses to clock time in some cases.[49] Also in Kenya, for data collection with men, limitations include that only one data collector was present (which may have impacted data quality) and that the sample of men was older on average than the sample of women (which may mean that we missed younger men who may perform different activities). And as with all time use surveys that use an interview-recall method, data may be prone to measurement error due to recall bias. However, we used multiple strategies for quality control, including recording and transcribing all interviews, which allowed us to check the accuracy of our data.

Another strength of our study is that we incorporated the concept of time-use agency and tested survey items to measure time-use agency, which can be used in future research in Kenya. However, due to translation and comprehension issues with the time-use agency scale in Honduras, we were unable to use the data from that setting. In addition, our sample size for factor analysis of the time-use agency items was modest; we acknowledge that “larger samples increase the generalizability of the conclusions reached by means of factor analysis”[29] and recommend future replication of this work with larger samples to demonstrate its generalizability.

## Conclusions

Our work makes several contributions to the literature, including by collecting data on women’s and men’s activities *including but not limited to* water collection, thereby filling gaps in standard time-use surveys and in surveys focused only on water collection. We also add to the evidence on potential effects of infrastructure projects for women’s time use and on the measurement of the relatively new concept of time-use agency. Our results suggest that more transformative programs and policies are needed to effectively address the needs of women and their households and to bring about meaningful change in women’s lives. More specifically, water interventions should provide household-level piped water that is continuously available in sufficient quality and quantity to meet all household needs year-round and should also include program components that aim to transform gendered social norms related to water work. Our time-use agency results from Kenya suggest that with appropriate infrastructural improvements, women may indeed have the decision-making power to reallocate their time as they choose. At the global level, to achieve SDG 5 and specifically Target 5.4, as well as SDG Target 6.1, more work must be done to continue to provide water infrastructure that meets all household needs and to transform gender norms.

## Data Availability

All data produced in the present study are available upon reasonable request to the authors.

## ACKNOWLEDGEMENTS

We are deeply grateful to the participants for their time and for sharing their experiences. We also extend our thanks to our partners and data collection teams whose collaboration made this research possible. From the Honduras research team, we thank Gladys Rosario Alvarez Montoya, Ruth Avigail Montalban, Ana Carolina Padilla, Luis Antonio Zelaya, Marta Ramos, Pedro Martinez, Salomon Perdomo, Sobeyda Rodriguez, Varinia Dominguez, Mayra Portillo, Maira Velasquez, Luisa Galindo, and Hernan Lanza. From the World Vision Honduras team, we thank Jazmina Nohemí Irías, Ana Lili Burgoos, and Carlos Parrales. From the Kenya research team, we thank Alois Benedicto, Abel Senaika, Andrew Solomon, Nasieku Lolchura, Patricia Lekilito, Lekalantula Raina Deborah, Iliano Loldepe, Longonyek Felista, Ntusunye Leliko, Jonathan Letipo, Naanyu Leliko, Gloria Karimi Kiriinya, Lekonte Salome Mary, and Ruth Kilimo. From the World Vision Kenya team, we thank Everlyne Atandi, Beatrice Mwai, and Kevin Muche. Lastly, we would like to thank Jorge Beteta for his translation services.

## SUPPORTING INFORMATION CAPTIONS

**Table S1**. Time Use Module

**Table S2**. Time-Use Agency Module

**Figure S1**. Average self-reported time use in hours among only those participants who reported participating in an activity, in Kenya sample (N=95), by activity type and gender

**Figure S2**. Mean time spent on seven broad activity categories during a 24-hour period among all women and men in Kenya, by gender (N = 95)

**Figure S3**. Self-reported participation in seven broad activity categories over the course of a 24-hour data collection period, by gender, Kenya sample (N=95)

**Figure S4**. Mean time spent on seven broad activity categories during a 24-hour period among all women in Kenya, by water source provision (N = 48)

**Figure S5**. Average self-reported time use in hours among only those participants who reported participating in an activity, in Honduras sample (N=102), by activity type and gender

**Figure S6**. Mean time spent on seven broad activity categories during a 24-hour period among all women and men in Honduras, by gender (N = 102)

**Figure S7**. Self-reported participation in seven broad activity categories over the course of a 24-hour data collection period, by gender, Honduras sample (N=102)

**Figure S8**. Mean time spent on seven broad activity categories during a 24-hour period among all women in Honduras, by water source provision (N = 102)

**Table S3**. Responses to items designed to measure time-use agency, Kenya sample (N=95)

## Notes

### Competing Interest Statement

The authors have declared no competing interest.

### Author Declarations

Ethical approvals were provided by the institutional review boards of Emory University (Atlanta, GA: STUDY00005955), Universidad Nacional Autonoma de Honduras (Tegucigalpa: CEIFCS-2023-P18), and St. Paul University (Nairobi: SPU/29/2023). A research license was granted by Kenya's National Commission for Science, Technology, and Innovation (Nairobi: NACOSTI/P/24/3752).

