## Supplemental Materials for "Improved water access may not reduce women’s time burdens: Evidence from Kenya and Honduras"

**Table S1. Time Use Module**

Now I’d like to ask you about how you spent your time during the past 24 hours. We’ll begin from yesterday morning, and continue through to this morning. This will be a detailed accounting. I’m interested in everything you did (i.e. resting, eating, personal care, work inside and outside the home, caring for children, cooking, shopping, socializing, etc.), even if it didn’t take you much time.

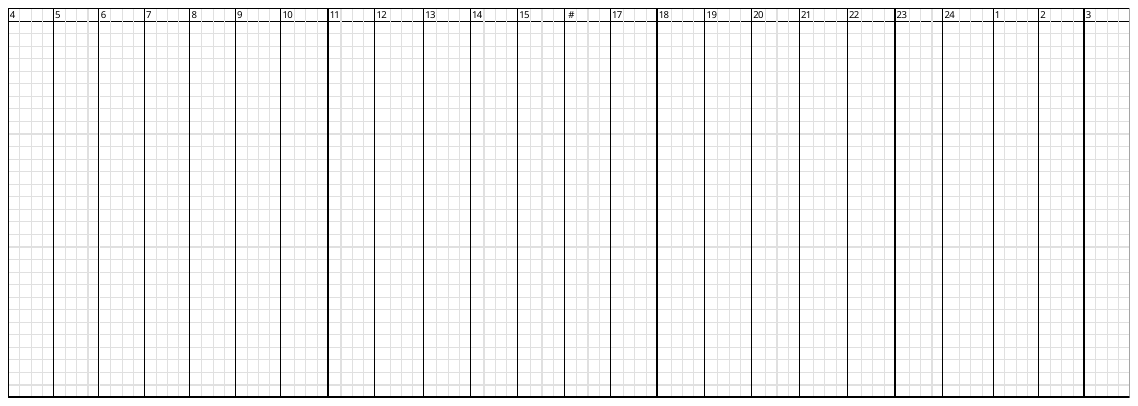

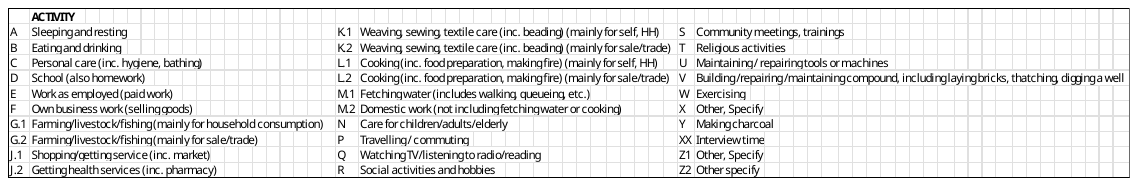

**Table S2. Time-Use Agency Module**

| 1. **Intrinsic Agency (Self-efficacy)** | |
| --- | --- |
| **INTERVIEWER, READ ALOUD:** Now, I am going to read several statements of **HOW SURE YOU FEEL THAT YOU CAN DO CERTAIN THINGS**. Please listen to each statement and **indicate whether you agree or disagree with the statement.**. | |
| You feel that you can **change your daily schedule**.  Do you completely or partly [agree/disagree]? | 0 = Completely disagree  1 = Partly disagree  2 = Partly agree  3 = Completely agree  88 = Refuse 66 = Not applicable 99 = Cannot answer |
| You feel that you can **ask a household member** to **do some of your household duties**.  Do you completely or partly [agree/disagree]? | 0 = Completely disagree  1 = Partly disagree  2 = Partly agree  3 = Completely agree  88 = Refuse 66 = Not applicable 99 = Cannot answer |
| You feel that you can **ask a household member** to **help you take care of a child or other family member**.  Do you completely or partly [agree/disagree]? | 0 = Completely disagree  1 = Partly disagree  2 = Partly agree  3 = Completely agree  88 = Refuse 66 = Not applicable 99 = Cannot answer |
| You feel that you can **change the amount of time you spend on paid work**.  Do you completely or partly [agree/disagree]? | 0 = Completely disagree  1 = Partly disagree  2 = Partly agree  3 = Completely agree  88 = Refuse 66 = Not applicable 99 = Cannot answer |
| 2. **Instrumental Agency (Decision-making)** | |
| **INTERVIEWER, READ ALOUD:** Now I am going to ask you about how much influence you have in decisions about your time. For each activity, please rate **THE AMOUNT OF INFLUENCE YOU HAD OVER TIME YOU SPENT ON THE ACTIVITY DURING THE LAST FOUR WEEKS.** If you did NOT do the activity, please think about how much influence you had over the decision to NOT participate. Please indicate whether you had **no influence, some influence, or a lot of influence** regarding how you spent your time.  **NOTE TO INTERVIEWER: When examples are given, they should be read exactly as written and should NOT be skipped.** | |
| **Prompt** | **Answer Choices** |
| During the last 4 weeks, how much influence did you have in decisions about the amount of time you spent on the following activities: |  |
| A. Household duties, such as cooking, cleaning, washing clothes, or collecting water or cooking fuel | 1 = No influence  2 = A small amount of influence  3 = A medium amount of influence  4 = A lot of / high influence  99 = DO NOT READ: Don’t know  88 = DO NOT READ: Refused |
| B. Caring for household members, such as children or older family members | 1 = No influence  2 = A small amount of influence  3 = A medium amount of influence  4 = A lot of / high influence  99 = DO NOT READ: Don’t know  88 = DO NOT READ: Refused  66 = DO NOT READ: Not applicable |
| C. Going to the market to purchase essential items | 1 = No influence  2 = A small amount of influence  3 = A medium amount of influence  4 = A lot of / high influence  99 = DO NOT READ: Don’t know  88 = DO NOT READ: Refused  66 = DO NOT READ: Not applicable |
| D. Non-agricultural work activities, including: doing any activity to earn income or in-kind payment; helping in a family business; selling goods, including from farming or livestock rearing | 1 = No influence  2 = A small amount of influence  3 = A medium amount of influence  4 = A lot of / high influence  99 = DO NOT READ: Don’t know  88 = DO NOT READ: Refused |
| E. Commercial agriculture (in fields/farms), including: work on family farming, fishing, or livestock activities whose products are mainly for sale | 1 = No influence  2 = A small amount of influence  3 = A medium amount of influence  4 = A lot of / high influence  99 = DO NOT READ: Don’t know  88 = DO NOT READ: Refused  66 = DO NOT READ: Not applicable |
| F. Household agriculture (in backyards/homesteads), including: including: farming, fishing, or livestock activities in and around your homestead whose products are mainly for household consumption | 1 = No influence  2 = A small amount of influence  3 = A medium amount of influence  4 = A lot of / high influence  99 = DO NOT READ: Don’t know  88 = DO NOT READ: Refused  66 = DO NOT READ: Not applicable |
| G. Attending a community meeting | 1 = No influence  2 = A small amount of influence  3 = A medium amount of influence  4 = A lot of / high influence  99 = DO NOT READ: Don’t know  88 = DO NOT READ: Refused  66 = DO NOT READ: Not applicable |
| H. Sleeping or resting | 1 = No influence  2 = A small amount of influence  3 = A medium amount of influence  4 = A lot of / high influence  99 = DO NOT READ: Don’t know  88 = DO NOT READ: Refused |
| During the last 7 days, how much influence did you have in decisions about your daily schedule?  This may include the amount of time you spend on activities as well as when during the day or in what order you do activities. | 1 = No influence  2 = A small amount of influence  3 = A medium amount of influence  4 = A lot of / high influence  99 = DO NOT READ: Don’t know  88 = DO NOT READ: Refused |

**Figure S1.** Average self-reported time use in hours among only those participants who reported participating in an activity, in Kenya sample (N=95), by activity type and gender

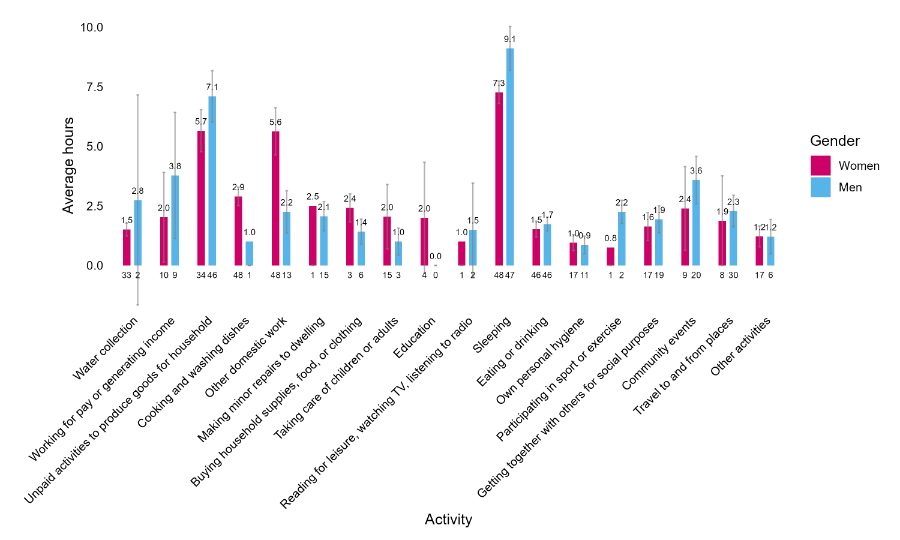
***Note:*** Numbers below bars indicate the number of respondents; numbers above bars indicate the average time spent on the activity

**Figure S2.** Mean time spent on seven broad activity categories during a 24-hour period among all women and men in Kenya, by gender (N = 95)

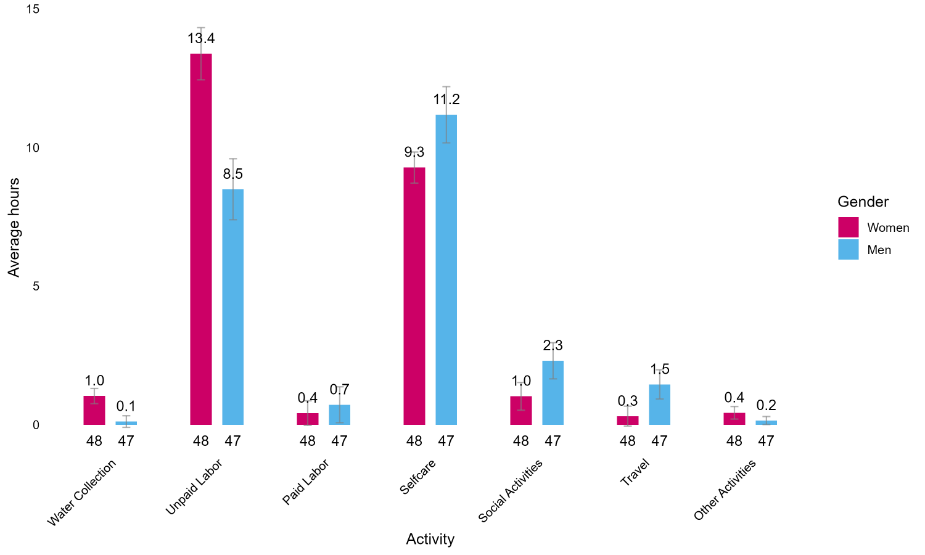

***Note:*** Numbers below bars indicate the number of respondents; numbers above bars indicate the average time spent on the activity

**Figure S3.** Self-reported participation in seven broad activity categories over the course of a 24-hour data collection period, by gender, Kenya sample (N=95)
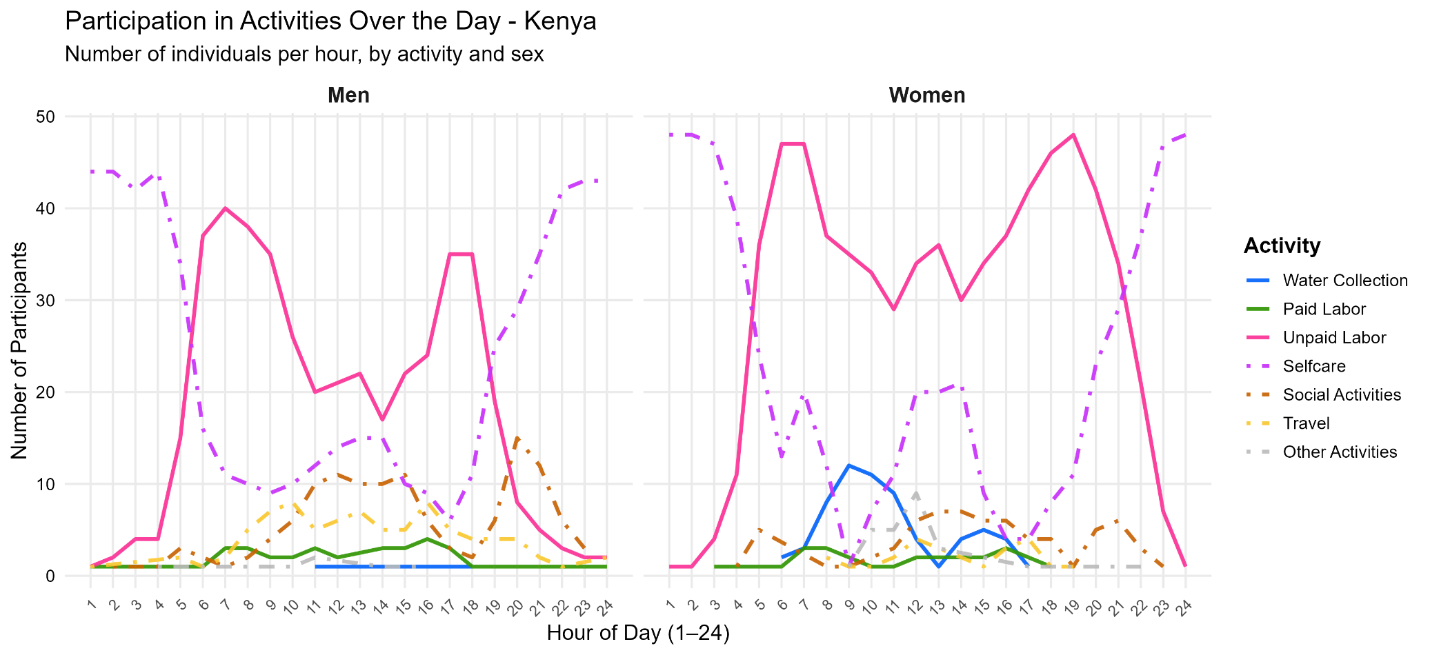

**Figure S4.** Mean time spent on seven broad activity categories during a 24-hour period among all women in Kenya, by water source provision (N = 48)

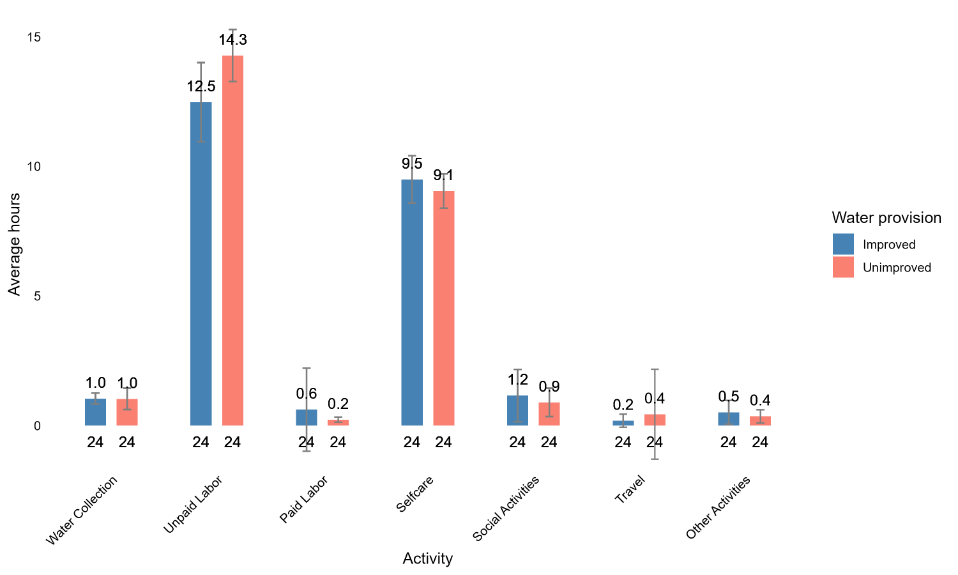

**Figure S5.** Average self-reported time use in hours among only those participants who reported participating in an activity, in Honduras sample (N=102), by activity type and gender

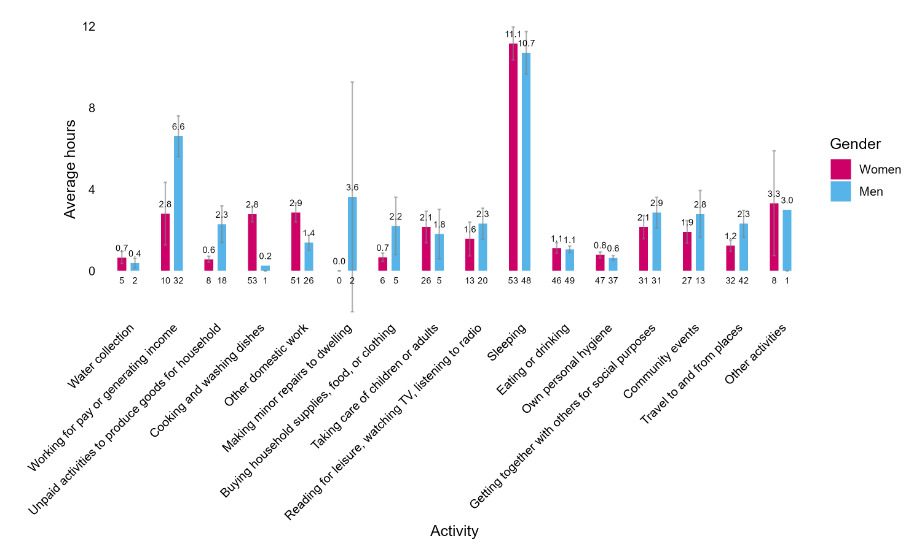

***Note:*** Numbers below bars indicate the number of respondents; numbers above bars indicate the average time spent on the activity

**Figure S6.** Mean time spent on seven broad activity categories during a 24-hour period among all women and men in Honduras, by gender (N = 102)

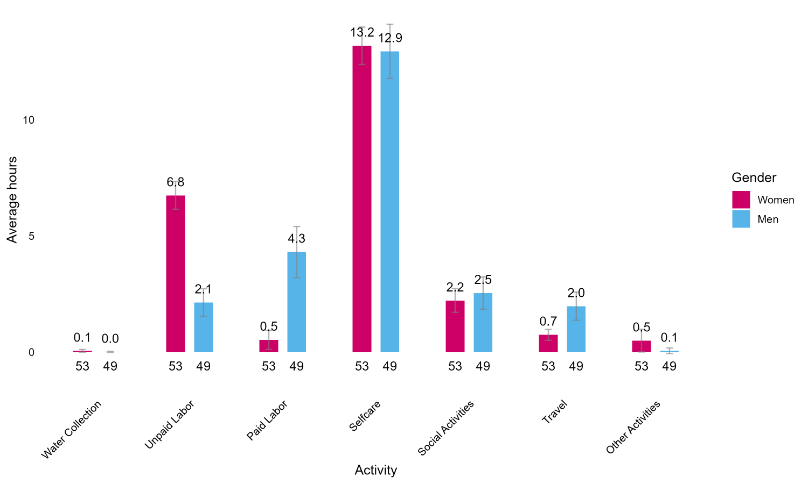

***Note:*** Numbers below bars indicate the number of respondents; numbers above bars indicate the average time spent on the activity

**Figure S7.** Self-reported participation in seven broad activity categories over the course of a 24-hour data collection period, by gender, Honduras sample (N=102)

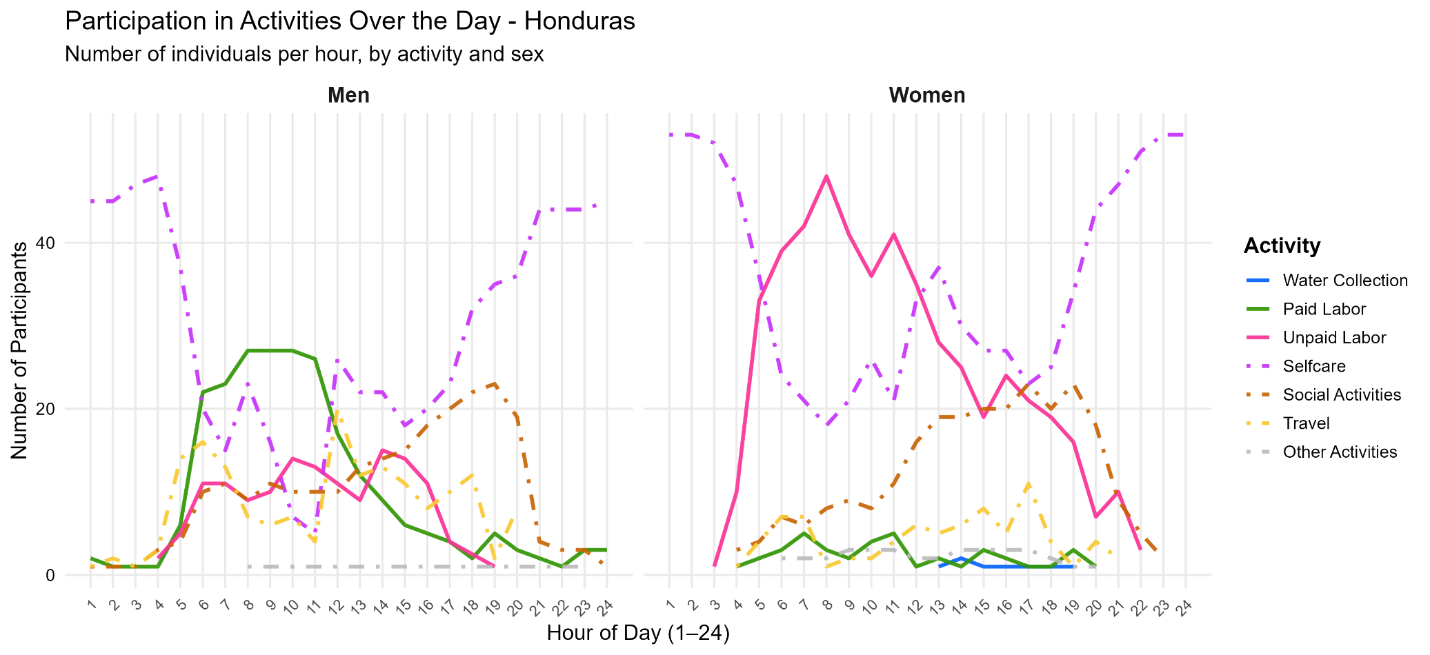

**Figure S8.** Mean time spent on seven broad activity categories during a 24-hour period among all women in Honduras, by water source provision (N = 102)

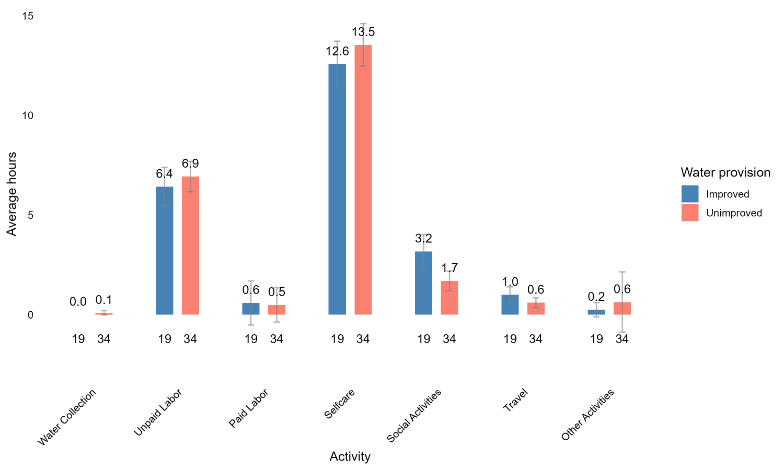

***Note:*** Numbers below bars indicate the number of respondents; numbers above bars indicate the average time spent on the activity

**Table S3.** Responses to items designed to measure time-use agency, Kenya sample (N=95)

|  | **Men** | | **Women** | |
| --- | --- | --- | --- | --- |
|  | **N** | **%** | **N** | **%** |
| **Intrinsic time-use agency (Self-efficacy related to time use)** |  |  |  |  |
| **You feel that you can change your daily schedule.** | 46 |  | 46 |  |
| Completely disagree | 3 | 6.52 | 3 | 6.52 |
| Partly disagree | 2 | 4.35 | 0 | 0 |
| Partly agree | 6 | 13.04 | 8 | 17.39 |
| Completely agree | 35 | 76.09 | 35 | 76.09 |
| **You feel that you can ask a household member to do some of your household duties.** | 46 |  | 47 |  |
| Completely disagree | 0 | 0 | 3 | 6.38 |
| Partly disagree | 1 | 2.17 | 0 | 0 |
| Partly agree | 3 | 6.52 | 6 | 12.77 |
| Completely agree | 42 | 91.3 | 38 | 80.85 |
| **You feel that you can ask a household member to help you take care of a child or other family member.** | 46 |  | 46 |  |
| Completely disagree | 3 | 6.52 | 0 | 0 |
| Partly disagree | 0 | 0 | 1 | 2.17 |
| Partly agree | 6 | 13.04 | 2 | 4.35 |
| Completely agree | 37 | 80.43 | 43 | 93.48 |
| **You feel that you can change the amount of time you spend on paid work.** | 46 |  | 43 |  |
| Completely disagree | 10 | 21.74 | 3 | 6.98 |
| Partly disagree | 1 | 2.17 | 0 | 0 |
| Partly agree | 1 | 2.17 | 3 | 6.98 |
| Completely agree | 34 | 73.91 | 37 | 86.05 |
| **Instrumental time-use agency (Decision-making / Influence over time use)** |  |  |  |  |
| **During the last 4 weeks, how much influence did you have in decisions about the amount of time you spent on the following activities:** | | | | |
| **Household duties, such as cooking, cleaning, washing clothes, or collecting water or cooking fuel** | 46 |  | 47 |  |
| No influence | 9 | 19.57 | 0 | 0 |
| A small amount of influence | 11 | 23.91 | 0 | 0 |
| A medium amount of influence | 6 | 13.04 | 1 | 2.13 |
| A lot of / high influence | 20 | 43.48 | 46 | 97.87 |
| **Caring for household members, such as children or older family members** | 46 |  | 47 |  |
| No influence | 3 | 6.52 | 0 | 0 |
| A small amount of influence | 1 | 2.17 | 0 | 0 |
| A medium amount of influence | 6 | 13.04 | 3 | 6.38 |
| A lot of / high influence | 36 | 78.26 | 44 | 93.62 |
| **Going to the market to purchase essential items** | 46 |  | 47 |  |
| No influence | 3 | 6.52 | 1 | 2.13 |
| A small amount of influence | 0 | 0 | 3 | 6.38 |
| A medium amount of influence | 9 | 19.57 | 10 | 21.28 |
| A lot of / high influence | 34 | 73.91 | 33 | 70.21 |
| **Non-agricultural work activities, including doing any activity to earn income or in-kind payment; helping in a family business; selling goods, including from farming or livestock rearing** | 46 |  | 47 |  |
| No influence | 2 | 4.35 | 6 | 12.77 |
| A small amount of influence | 0 | 0 | 5 | 10.64 |
| A medium amount of influence | 8 | 17.39 | 6 | 12.77 |
| A lot of / high influence | 36 | 78.26 | 30 | 63.83 |
| **Commercial agriculture (in fields/farms), including work on family farming, fishing, or livestock activities whose products are mainly for sale** | 43 |  | 45 |  |
| No influence | 15 | 34.88 | 7 | 15.56 |
| A small amount of influence | 0 | 0 | 4 | 8.89 |
| A medium amount of influence | 6 | 13.95 | 6 | 13.33 |
| A lot of / high influence | 22 | 51.16 | 28 | 62.22 |
| **Household agriculture (in backyards/homesteads), including including: farming, fishing, or livestock activities in and around your homestead whose products are mainly for household consumption** | 46 |  | 47 |  |
| No influence | 5 | 10.87 | 1 | 2.13 |
| A small amount of influence | 2 | 4.35 | 3 | 6.38 |
| A medium amount of influence | 11 | 23.91 | 6 | 12.77 |
| A lot of / high influence | 28 | 60.87 | 37 | 78.72 |
| **Attending a community meeting** | 45 |  | 45 |  |
| No influence | 1 | 2.22 | 0 | 0 |
| A small amount of influence | 1 | 2.22 | 6 | 13.33 |
| A medium amount of influence | 6 | 13.33 | 10 | 22.22 |
| A lot of / high influence | 37 | 82.22 | 29 | 64.44 |
| **Sleeping or resting** | 46 |  | 47 |  |
| No influence | 0 | 0 | 2 | 4.26 |
| A small amount of influence | 4 | 8.7 | 2 | 4.26 |
| A medium amount of influence | 9 | 19.57 | 1 | 2.13 |
| A lot of / high influence | 33 | 71.74 | 42 | 89.36 |
| **During the last 7 days, how much influence did you have in decisions about your daily schedule?** | 46 |  | 47 |  |
| No influence | 0 | 0 | 0 | 0 |
| A small amount of influence | 0 | 0 | 3 | 6.38 |
| A medium amount of influence | 9 | 19.57 | 4 | 8.51 |
| A lot of / high influence | 37 | 80.43 | 40 | 85.11 |
